# The SARS-CoV-2 epidemic outbreak: a review of plausible scenarios of containment and mitigation for Mexico

**DOI:** 10.1101/2020.03.28.20046276

**Authors:** Manuel Adrian Acuña-Zegarra, Andreu Comas-García, Esteban Hernández-Vargas, Mario Santana-Cibrian, Jorge X. Velasco-Hernandez

## Abstract

We present here several variants of a mathematical model to explore three main issues related to SARS-CoV-2 spread in scenarios similar to those present in Mexico and elsewhere in Latin America. We explore the consequences for travel inside a given region, in this case Mexico, particularly focusing on airplane transportation but attempting to give a gross approximation to terrestrial movement since this is the main form of population movement across geographical areas in the country; then we proceed to study the effect of behavioral changes required to lower transmission by lowering the contact rate and infection probability and lastly, we explore the consequences of disease spread in a population subject to social isolation.These models are not suitable for predictive purposes although some rough predictions can be extracted from them. They are presented as a tool that can serve to explore plausible scenarios of spread and impact, effectiveness and consequences of contention and mitigation policies. Given the early stage at which the epidemic is at the date of writing in Mexico, we hope these ideas can be helpful for the understanding of the importance of isolation, social distancing and screening of the general population.

**Key findings:**

- We have estimated the parameters of the epidemic curve (growth rate, carrying capacity and dispersion) as well as a first estimate of the basic reproduction number for Mexico.
- We provide expected trends of epidemic outbreaks depending upon of the number of imported cases per day arriving to a large airport. We illustrate this trends with data from Mexico City airport.
- We provide expected trends of disease dispersal depending upon of the number of exported cases per day either by airplane or bus. We illustrate this trends with data from Mexico City.
- We evaluate the effect of behavioral change to reduce the contact rate and compare diverse scenarios that evaluate the timing of initial enforcement of behavior, time horizon in which to diminish the contact rate and the proportion of people under isolation.
- We evaluate the effect of social isolation by itself with respect to two main parameters: the starting time for the enforcement of control measures, and the learning time to achieve the desired contact rate reduction. We stress the importance of quick and direct actions to isolate and reduce contact rate simultaneously.

## 1 Introduction

At the end of December of 2019 a novel coronavirus SARS-CoV-2, was detected in adults with pneumonia from the city of Wuhan, Hubei Province, China. This new virus rapidly spread from Hubei Province to all China [1–5]. By January 22th the virus was detected in South Korea, Taiwan, Thailand. Macao and USA. By March 12 of 2020, 128,343 cases of new virus have been detected in 116 countries and 63% of the cases has been register in Continental China. In February 26, Brazil was the first country of Latin America to detect this virus and Mexico detect his first case one day later [6]. CoVID-19 is a respiratory disease caused by the SARS-CoV-2 virus, that has been widely studied in these recents weeks and we defer its description to the appropriate sources e.g. [4, 5]. The disease has been extensively modeled as a SEIR type of epidemic. In this work we use the SEIR model framework adapting it to the particular set of problems addressed here. In each section, dependent on the model variant used, we introduce the new parameters if any and refer to the appropriate section in the Appendix for the specific model equations. *The rate ω* or the related time 1/*ω* is the mean time of application of non-pharmaceutical interventions (NPI). All the recommendations that are being advised to prevent infection affect behavior (washing hands, keeping cough/sneeze etiquette, avoiding handshakes, keeping distance from other people) and, therefore, have a learning curve and can be forgotten or relaxed or incompletely adopted in any given population; also *q* is the proportion of the population that is effectively put under isolation. For example, in Wuhan, about 60% of the population was put under isolation. The total population normally living in that region is of about 14 million but the population at risk was obviously larger comprising much of the Hubei region.

The incubation and infectious periods have been a most important issue in the present epidemic and estimates have varied as more information is available on the dynamics of the disease. We will use incubation period 1/*γ* = 6 days [3, 7–9]. The same situation occurs with the infectious period that we take 1/*η* = 10 days [7, 10, 11]. One of the first articles to calculate the basic reproduction number *R*_0_ of the new coronavirus in Wuhan China, estimated (Jan 24, 2020) its value as 2.68(2.47 − 2.86) with an epidemic doubling time of 6.4 days (5.8 − 7.1) [3]. Other authors have estimated the basic reproductive number to be around 2.5, value that we use here for the Wuhan original outbreak but again there exist some variability and uncertainity in the different estimates [9–14]. In what follows some simple models are described related to three specific situations described above and occurring during this epidemic: disease dispersion due to long range population movement, behavioral change imposed on individuals as an strategy of containment, and the effect of social distancing on the general fate of the epidemic.

### 2 Intial growth: where Mexico stands

We analyze the cumulative trajectory of COVID19 cases in Mexico in order to create short-term real-time forecasts of the outbreak. The data consists of daily cumulative cases from February 28, 2020 to March 26, 2020. The data was obtained daily at 19:00 p.m. (GMT-6) from the Secretariat of Health press conferences.

To create the forecasts, we use Richards model which considers that cumulative cases *C*(*t*) are given by

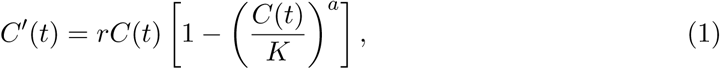

where *r* represents the growth rate of infection, *K* is the carrying capacity (or final epidemic size, and *a* is a scaling parameter. This model is an extension of the simple logistic growth model that has been recently used to predict cumulative COVID19 cases in China [15]. Parameters *a, r* and *K* must be estimated in order to properly describe the observed data and to provide accurate forecasts. We use an statistical approach through Bayesian inference. Details regarding the estimation procedure and the the results can be found on Appendix A. The growth rate *r* is close to 0.2 and it can range from 1.9 to 2.1 with a probability of 95%. However, the total size of the outbreak *K* is more difficult to estimate which is expected since Mexico is at the start of the outbreak and the current data shows that the contagion is accelerating. We estimate the final size of the outbreak to be 710,392 cases, ranging from 65,163 up to 1,258,296 with a probability of 95% provided no cotrol/contaiment/mitigation measures are applied. These are very long-term predictions and that is why there is too much variability in the interval. To estimate the parameters we used data up to March 24, 2020. The last two available observations corresponding to March 25 and 26 can be seen in Figure 1, and are closely predicted by the fitted model. Furthermore, we create 14 days predictions from March 25, 2020 to April 7, 2020 which can also be seen in Figure 1. For example, at March 31, 2020, the model expects 1,103 cases, with a 95% probability interval of (401, 2353); and at April 7, 2020, a total of 4,202 cases are expected with a 95% probability interval of (1,440, 11,012). Table 1 shows the median posterior estimates for each parameter and 95% probability intervals. Figure 2 shows the marginal posterior distributions estimates for each parameter.

**Table 1:**
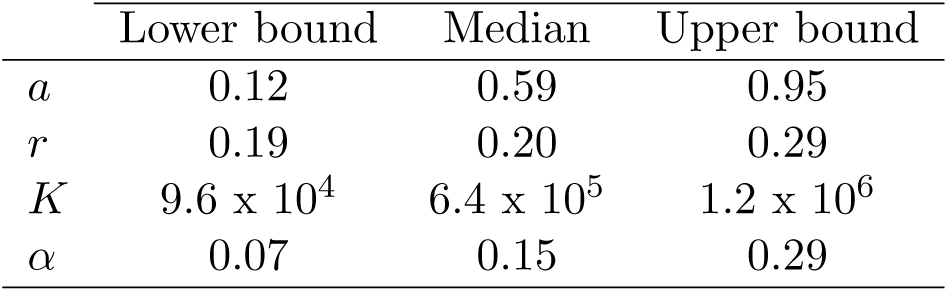
Posterior estimates for each parameter and 95% probability intervals. Only data from February 28,2020 to March 24, 2020 was used to produce these estimates, the last two available observations were removed.

|  | Lower bound | Median | Upper bound |
| --- | --- | --- | --- |
| $a$ | 0.12 | 0.59 | 0.95 |
| $r$ | 0.19 | 0.20 | 0.29 |
| $K$ | $9.6 \times 10^4$ | $6.4 \times 10^5$ | $1.2 \times 10^6$ |
| $\alpha$ | 0.07 | 0.15 | 0.29 |

**Figure 1:**
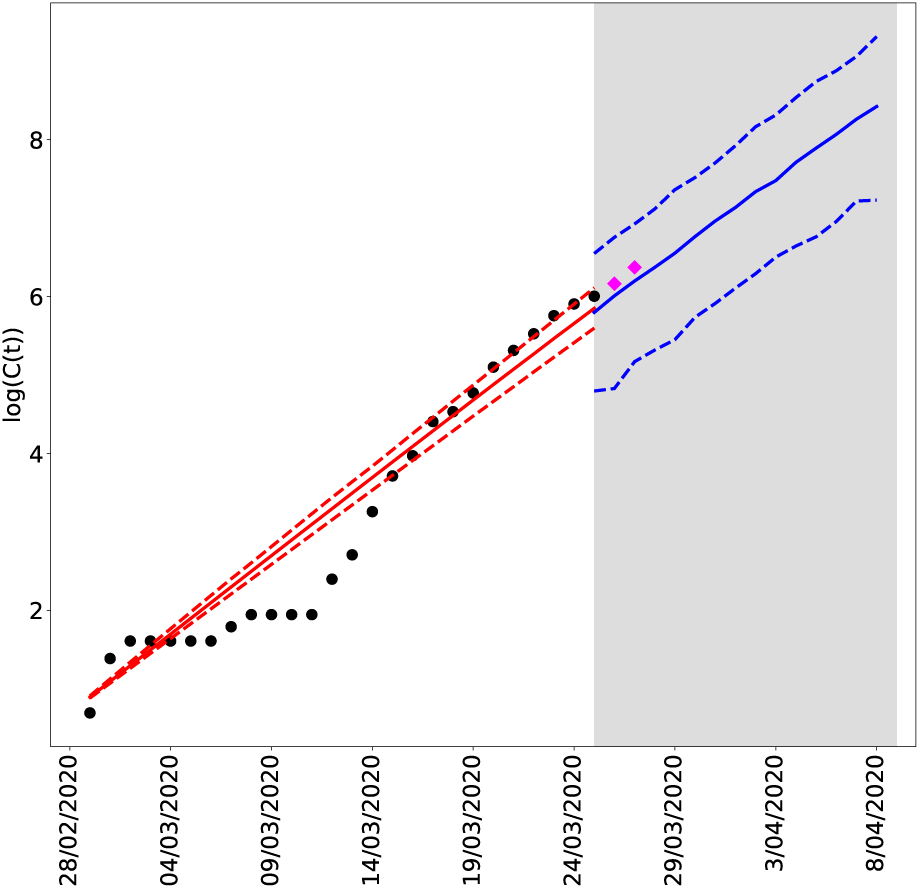
Cumulative COVID19 cases in Mexico from February 28, 2020 to March 26, 2020. Black dots show the observed cases. Pink diamonds are the two most recent observations that were not used to fit the model in order to test the short term accuracy of the model. The red solid line shows the mean trajectory of the outbreak estimated from the data, and red dashed lines are 95% probability intervals. The blue solid lines represent the mean posterior predictive trajectory of the epidemic and the blue dashed lines are 95% predictive probability intervals.

**Figure 2:**
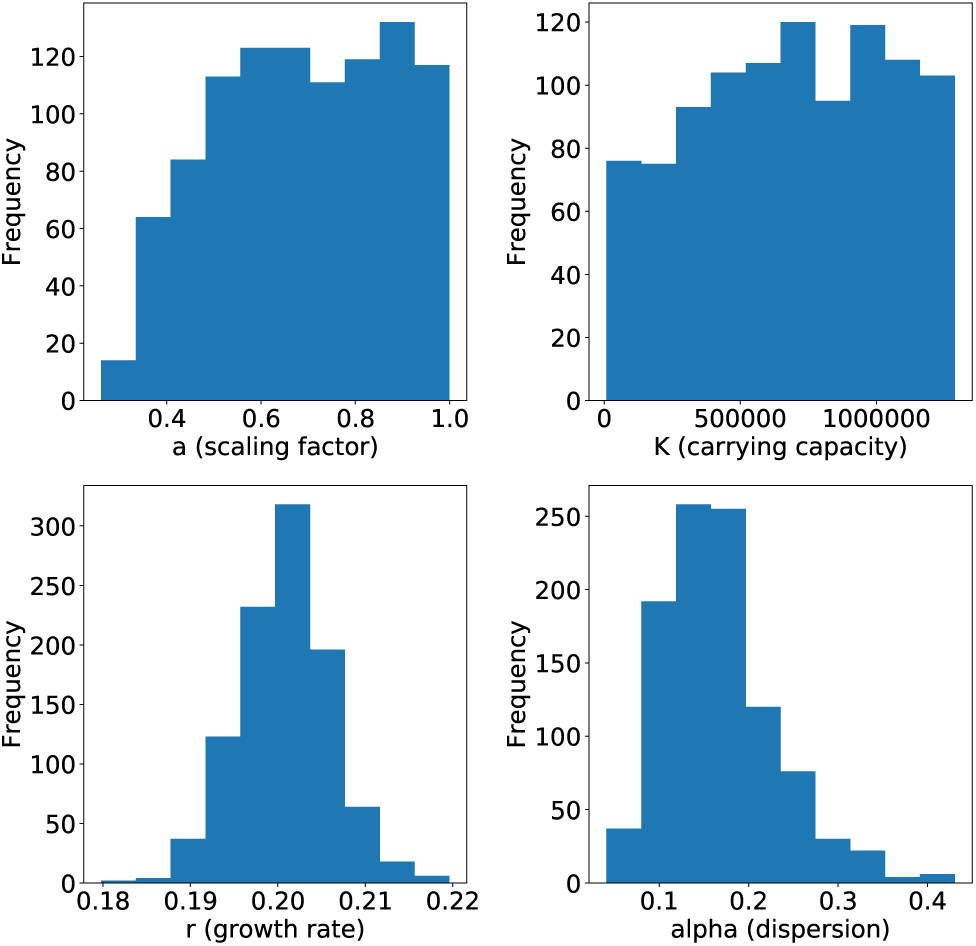
Posterior distributions for the scaling factor, carrying capacity, growth rate and dispersion of predicted cases

Finaly, using the same data we estimated the basic reproduction number for the whole country. We use the package written in R, Estimation of R0 and Real-Time Reproduction Number from Epidemics (https://CRAN.R-project.org/package=R0) using the exponential and maximum likelihood methods. We obtained using the exponential growth method *R*_0_ = 2.7 (2.5, 3.2); and using the maximum likelihood *R*_0_ = 2.3 (2.0, 2.6). Given that for the Richards curve, the growth rate for Mexico is *r* = 0.2 below of that of the Wuhan epidemic, see Table 1, we use the estimate from the maximum likelihood method that renders *R*_0_ = 2.3 with an uncertainity interval of (2.0, 2.6).

In what follows we use several mathematical models to explore the consequences of interventions on the spread of the disease, i.e., how interventions modify the worrisome picture depicted in Figure 1.

## 3 Dispersion by air and road transportation

The Mexico City airport is one of the main connections hubs of Latin America and the country, and therefore arrival of COVID19 cases has occurred. In this section, we use data on the daily number of domestic and international travelers that arrive/leave to/from the Mexico City airport [16]. Below we follow the ideas presented first in [3]. We construct an standard SEIR Kermack-McKendrick mathematical model (see Appendix B) where *s*, *e*, *y*_*a*_, *y*_*s*_ and *r* represent susceptible, exposed, asymptomatic infectious, symptomatic infectious, and recovered (immune) individuals. *N* is the total population. The force on infection is (*β*_*a*_*y*_*a*_ + *β*_*s*_*y*_*s*_)/*N*. We consider that symptomatic individuals have restricted movement since either they are too sick to travel or are immediately detected by the health services at the airport or elsewhere. This consequently decreases their overall contact rate *β*_*s*_ compared to that of an asymptomatic individual *β*_*a*_. Therefore *β*_*s*_ < *β*_*a*_ throughout this section. Following [3] we define two rates related to the inbound number of international air passengers to Mexico City (*L*_*im*_), and the inbound number of national of both airplane and terrestrial passengers to Mexico City (*L*_*sm*_). The basic model is shown in Appendix B

As the main objective in this section is to study the consequences of disease importation to a large Latin American city, we assume first that a proportion of the international air passengers arrive as disease carriers. We define *ρ*_*t*_ to be the proportion of international air passengers at time *t* who cannot transmit the disease, e.g., if *ρ*_*t*_ = 0.75 then 25% of the international air passengers are infectious. Of these carriers, a proportion 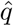 arrive as exposed not yet infectious individuals and 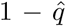 arrive as asymptomatic infectious individuals. *L*_*ms*_ and *L*_*sm*_ are functions of time describing travelers either arriving to Mexico City airport of leaving Mexico City airport. These rates are time dependent since we eventually take into account the increase on the number of national travelers during holiday periods, specifically Holy Week in Mexico. For the analysis we perform a normalization in order to work with proportions and not absolute numbers of individuals (see (6), Appendix C). One of the goals of this manuscript is to study the arrival of COVID19 to Mexico City and its plausible patterns of propagation. The timeline of our study is half a year. To estimate the daily average number of national and international inbound/outbound air and road travelers, respectively we employ information from the Secretariat of Communications and Transportation (SCT) [16] and the General Directorate for Tourism Competitiveness of Mexico City [17]. The timeline is set to include Holy Week, a short holiday which normally presents an increase of arrival/departure flights to/from Mexico City. On the other hand, the main transport means of the Mexican population is by far terrestrial by bus. Unfortunately, we only have data on the number of bus passengers outbound from Mexico City, reported by the large bus terminals Norte, Sur, Poniente and Oriente. We will assume that on average the number of bus passengers that arrive to Mexico City is greater than those that leave.

For our simulations, we employ flight data for February 2020, terrestrial data for January 2018/2019 and take February 21st as the initial date of the arrival of the first cases. This estimate is plausible since the first cases in Mexico were reported on February 28th but given that the incubation period is of 6 days they likely arrived to Mexico City on February 21st or 22nd. We also set the population size of the Greater Metropolitan Area of Mexico City at 25 210 748 inhabitants. To estimate the inbound/outbound *international* travelers migration rates, we take into account all flights that arrive/leave Mexico City [16]. Thus, the daily average number of international arrival and departure air travelers are *L*_*im*_ = 22, 519 and *L*_*mi*_ = 21, 838, respectively. See Appendix G for the procedure followed to give this estimates. To estimate the inbound/outbound *national* travelers migration rates we integrate air and terrestrial transportation. For national flights we arbitrarily select the thirteen more important destinations from Mexico City, i.e., those who have more passengers inbound leaving from Mexico City. On the other hand, to estimate the number of terrestrial passengers, we use data from the above mentioned four main bus terminals of Mexico City. Therefore, we calculate the averages per day of national passengers in both kinds of transport (airplane and bus). Thus *L*_*sm*_ = 141, 664 and *L*_*ms*_ = 141, 425 except during the Holy Week (for more details see Appendix G). For the 2020 Holy Week (April 4th to April 12th), we proceed to increase previous estimates by 25% which is a rough and conservative approximation based on data from the Secretariat of Tourism [17]. Therefore *L*_*sm*_ = 177, 080 and *L*_*ms*_ = 176, 781 during this period and also the information available on the main bus terminals of Mexico City (for further details see Appendix G). Finally, the incubation and the infectious period are set to *γ* = 1/6 and *η* = 1/10 per day, respectively. Also, since we do not have data on the air passenger percentage that arrives in the latent or asymptomatic stage, we propose as a plausible value 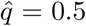 as a null hypothesis from which to start our analysis.

### 3.1 Case importation of asymptomatics

For our first case, we assume *ρ*_*t*_ = 0.999956 that is Mexico City airport received only one imported case per day (IPD). Figure 3 shows that the higher the proportion of asymptomatic infected arriving two things happen: the outbreak occurs earlier and also the peak of the outbreak is higher. In Table 2, we further illustrate this case. Notice that the changes in prevalence at different dates when proportions of asymptomatic infected is increased. For this model we explicitly calculated the basic reproductive number since it depends on *ρ* the proportion of asymptomatic infectious individuals, and explore the consequences of taking *ρ* = 0.30, 0.60 and 0.90, or, equivalently, ℛ_0_ = 2.0585, 2.1160 and 2.1719, respectively. These values are within the range reported by several authors and our own uncertainity interval (Section 1) [1, 10, 18].

**Table 2:**
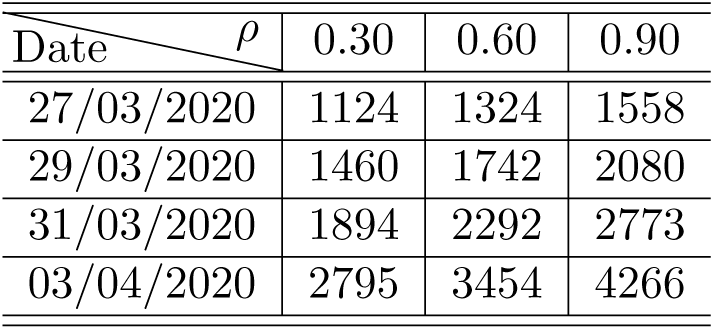
Mexico City prevalence (number of cases) for different days as a function of the percentage of asymptomatic cases, under the assumption of one IPD.

| Date \ $\rho$ | 0.30 | 0.60 | 0.90 |
| --- | --- | --- | --- |
| 27/03/2020 | 1124 | 1324 | 1558 |
| 29/03/2020 | 1460 | 1742 | 2080 |
| 31/03/2020 | 1894 | 2292 | 2773 |
| 03/04/2020 | 2795 | 3454 | 4266 |

**Figure 3:**
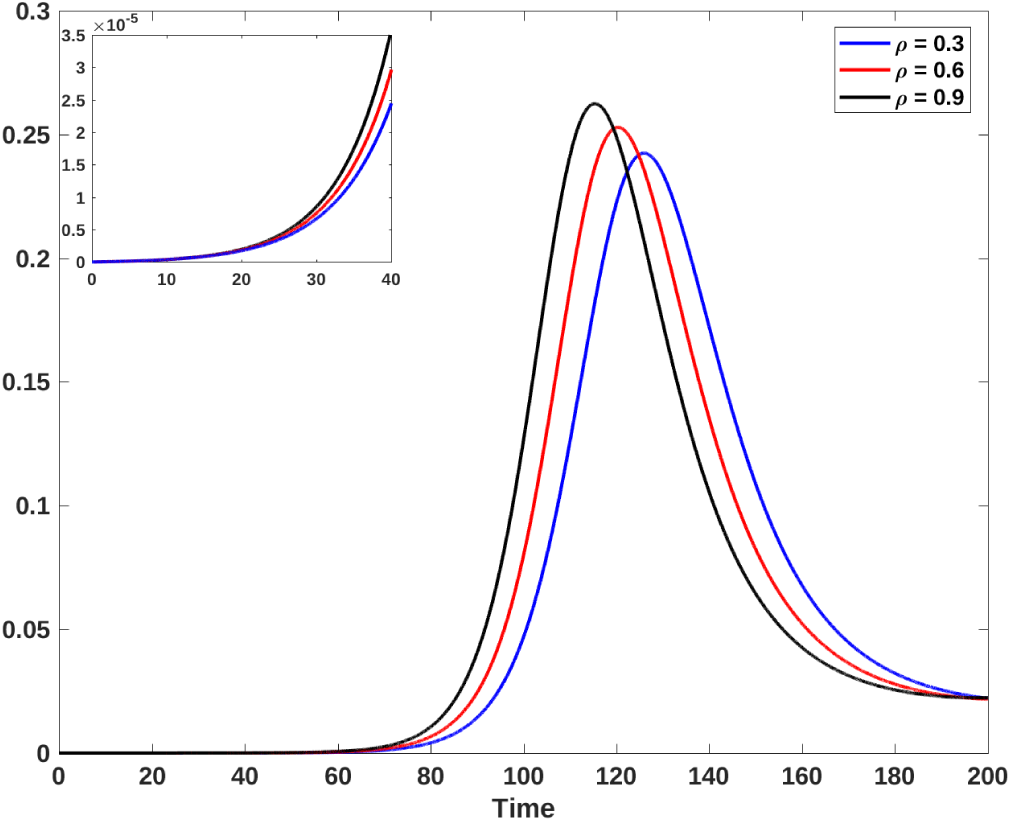
Prevalence for one IPD. Blue, red and black lines represent a 30%, 60% and 90% of asymptomatics, respectively. The upper left-hand panel shows the initial growth of the epidemic for each case. Horizontal axis is time in days and the vertical axis is proportions.

### 3.2 Varying number of imported cases

In our second case, we explore the effect of we explore the effect of decreasing *ρ*_*t*_ which is equivalent to increasing the number of IPD. Figure 4 shows that for a higher number of IPD generates an earlier outbreak but the maximum of the outbreak does not change. In Table 3, we further illustrate this case for Mexico City. Once again we observe that prevalence changes when increasing the number of IPD. This case is set by assuming that 80% (*ρ* = 0.8) of the infected people are asymptomatic giving ℛ_0_ = 2.1534.

**Table 3:** Mexico City prevalence (number of cases) for different days as a function of IPD (imported cases per day) under the assumption that 80% of cases end up as asymptomatic.

| Date \ IPD | 1 | 2 | 3 |
| --- | --- | --- | --- |
| 27/03/2020 | 1476 | 2952 | 4428 |
| 29/03/2020 | 1961 | 3921 | 5882 |
| 31/03/2020 | 2603 | 5206 | 7808 |
| 03/04/2020 | 3976 | 7953 | 11927 |

**Figure 4:**
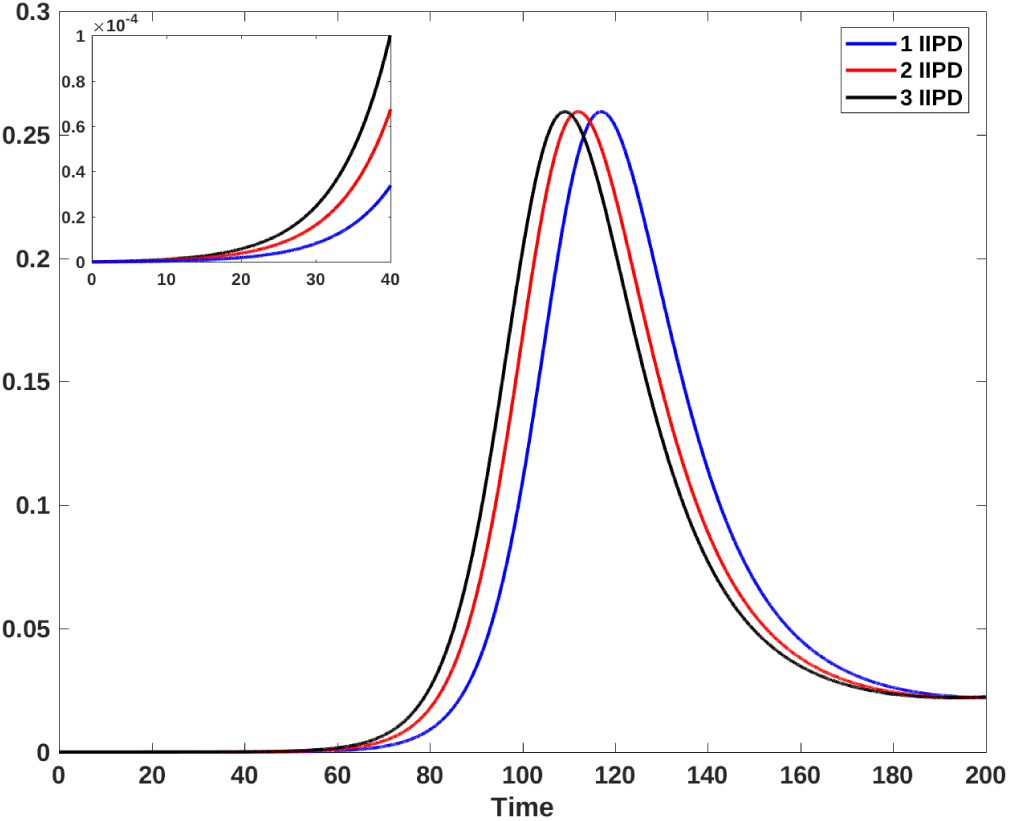
Prevalence for one (blue line), two (red line) and three (black line) IPD. The upper left-hand panel shows the initial growth of the epidemic for each case. Horizontal axis is time in days, vertical axis are proportions.

We remark that if we simulate both cases in (Figures 3-4) for a longer period of time, we can detect the existence of another epidemic outbreak as has already been noted [19].

### 3.3 Exportation from Mexico City

Figure 5 shows the expected number of exported cumulative cases (until April 1st) to several of the Mexican cities that normally receive the highest outbound air or road travel volumes. Here, as *ρ* = 0.8, ℛ_0_ = 2.1534. Figure 5(a) shows the expected number of exported cumulative cases that use air travel. When considering one IPD, we see that few states receive at least one exported case from Mexico City (Cancun, Guadalajara and Monterrey). On the other hand, with three IPD, these states import 4, 3 and 3 cases from Mexico City, respectively. Figure 5(b) shows the exported cumulative cases for terrestrial transportation. Here, if the imported cases per day (IPD) is one, then Puebla, Jalisco, Michoacan, Guerrero, and Queretaro import 4, 5, 3, 2 and 1 cases up to Aril 21st from Mexico City, respectively; while if IPD is equal to three, then Puebla, Jalisco, Michoacan, Guerrero, and Queretaro import 13, 16, 9, 7 and 4 infections from Mexico City, respectively.

**Figure 5:**
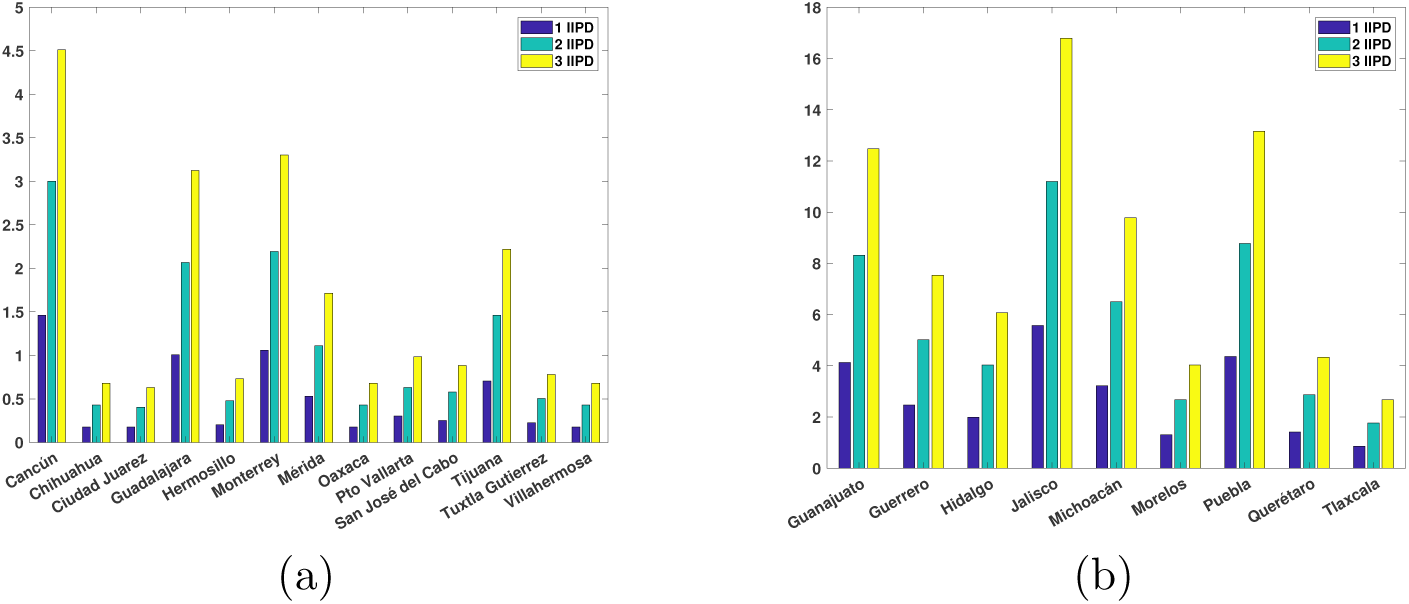
Expected number of cumulative cases exported to Mexican cities with the highest outbound travel volumes from Mexico City until April 1st. Air (*a*) and terrestrial (*b*) travel. These estimates correspond to one (blue bars), two (green bars) or three (yellow bars) IPD.

Figure 6 shows the expected number of exported cumulative cases to several Mexican cities. We simulate the number of cases until April 21st. We can observe the importance of the number of people who arrive infected since these can significantly increase the spread of the disease to other states. For example, Figure 6(a) shows that for Cancun, the number of exported cases can increase by 50 cases if instead of 1 IPD, we have 3 IPD. For Monterrey the increase, under the same conditions, is approximately 40 cases. Figure 6(b) shows a greater increase in cases for terrestrial than for air transportation. For example, we expect that for Jalisco the number of exported cases (by road transportation) can increase in 190 cases if instead of 1 IPD, we have 3 IPD. Similarly for Puebla where the number of exported cases can be increased in 145 cases approximately if instead 1 IPD, we have 3 IPD.

**Figure 6:**
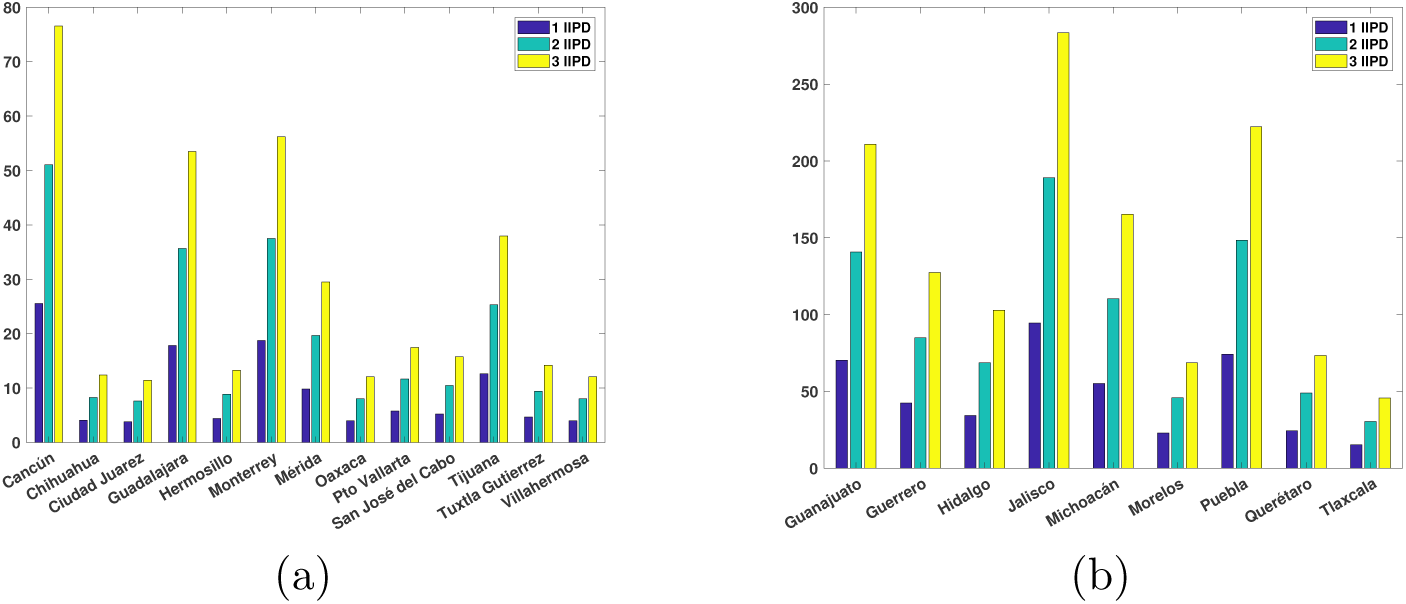
Expected number of cumulative cases exported to Mexican cities that have the highest outbound travel volumes from Mexico City until April 21st. Air (*a*) and terrestrial (*b*) travel. These estimates correspond to one (blue bars), two (green bars) or three (yellow bars) IPD.

Finally, it is an obvious conclusion from our simulations that it is very important to control the number of people who arrive infected since these can significantly increase the spread of the disease to the other states. Note that the results in this section do not include any control, mitigation of containment strategy. The results shown in Tables 2-3 are some possible scenarios in the *absence* of control interventions.

## 4 Behavioral change and epidemic containment

Once the disease arrives into a given region, city, or municipality, containment measures are at first, limited to behavioral changes called non-pharmaceutical interventions or NPIs, directed essentially to diminish transmission by reducing contact between people. In order to model this, we modify system (15) to include a variable contact rate 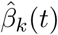, and omit migratory phenomena (see Appendix D for the model equations).

Our main results in this section explore the variation of the effective contact rate under these actions. We define

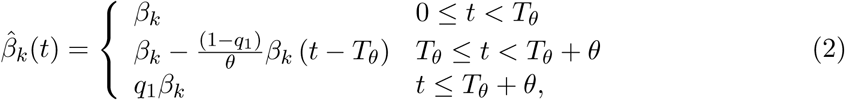

where *k* = *a* or *s* for asymptomatic or symptomatic individuals, respectively. Note from the formula above that we assume that behavioral change geared to diminish the contact rate from *β*_*k*_ to *q*_1_*β*_*k*_ takes a learning time *θ* to acquire where *q*_1_ is the desired proportion of reduction of the initial contact rate, e.g., *q*_1_ = 0.25 if the contact rate is reduced in 75% at the end of a time interval of length *θ*. Moreover, these control measures are not taken immediately after the outbreak is identified but lag a certain time *T*_*θ*_ before being enforced or (see Figure 7).

**Figure 7:**
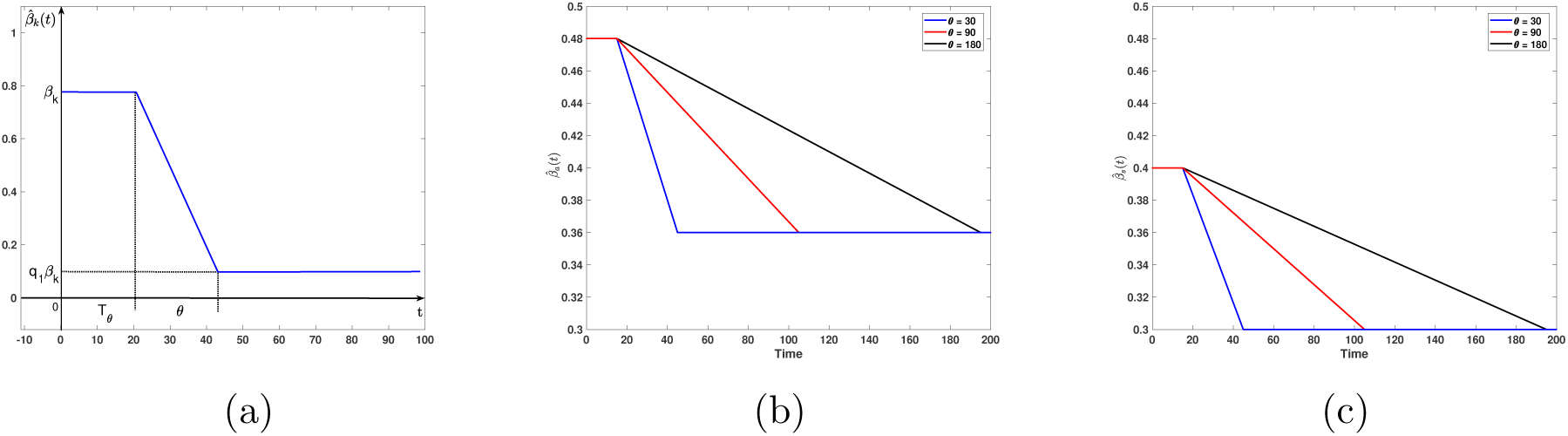
Time-dependent contact rates for a reduction. (*a*) General description of the contact rate parameters: *β*_*k*_ original contact rate, *q*_1_*β*_*k*_ target contact rate, *T*_*θ*_ time delay for the application of contact rate reduction, *θ* learning time to reduce contact rate to target level. (*b*) Contact rate reduction of 25% percent (*q*_1_ = 0.75) when *T*_*θ*_ = 15 for asymptomatic cases; (*c*) but same situation for the symptomatic cases. For (*b*) and (*c*) black, red and blue lines indicate times for contact rate reduction of *θ* = 180, 90 and 30 days, respectively.

### 4.1 Diminishing the contact rate

The baseline parameters for the simulations that follow are

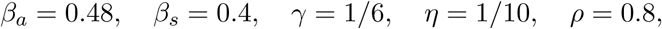

where, we remind the reader correspond to the contact rates of asymptomatic and symptomatic infecteds, the incubation and infectious period and the proportion of asymptomatic cases respectively.

Figure 7 illustrates the behavior of the contact rates for asymptomatics 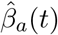 and for symptomatics 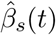. For both rates, the efficacy of education on the behavioral change is the same since we assume that education is not related to symptomatology although there is some evidence that this might not be the case. A first result is that if the time it takes to obtain the desired contact rate reduction *θ* increases then the reduction rate of both contact rates is slower.

Figures 8 and 9 show that if the time it takes to achieve a reduction in contact rate increases then the number of infectious cases is greater reflecting the slow rate of behavioral change. Figure 8 a) and b) show the prevalence and cumulative incidence respectively, for an scenario with a target reduction of contact rate of 25%, the initiation of the behavioral change is *T*_*θ*_ = 15 days after 10 cases are detected and where 80% of those are asymptomatic. For comparison, a red dashed line shows the epidemic curves when no behavioral change is enforced. Here we see that although the reduction of contact rate is low (25%) (*q*_1_ = 0.75), in the long run behavioral change helps to reduce the incidence. We also note that in addition to the reduction in cases, as the time to achieve the contact rate reduction increases, the epidemic outbreak starts earlier and is larger.

**Figure 8:**
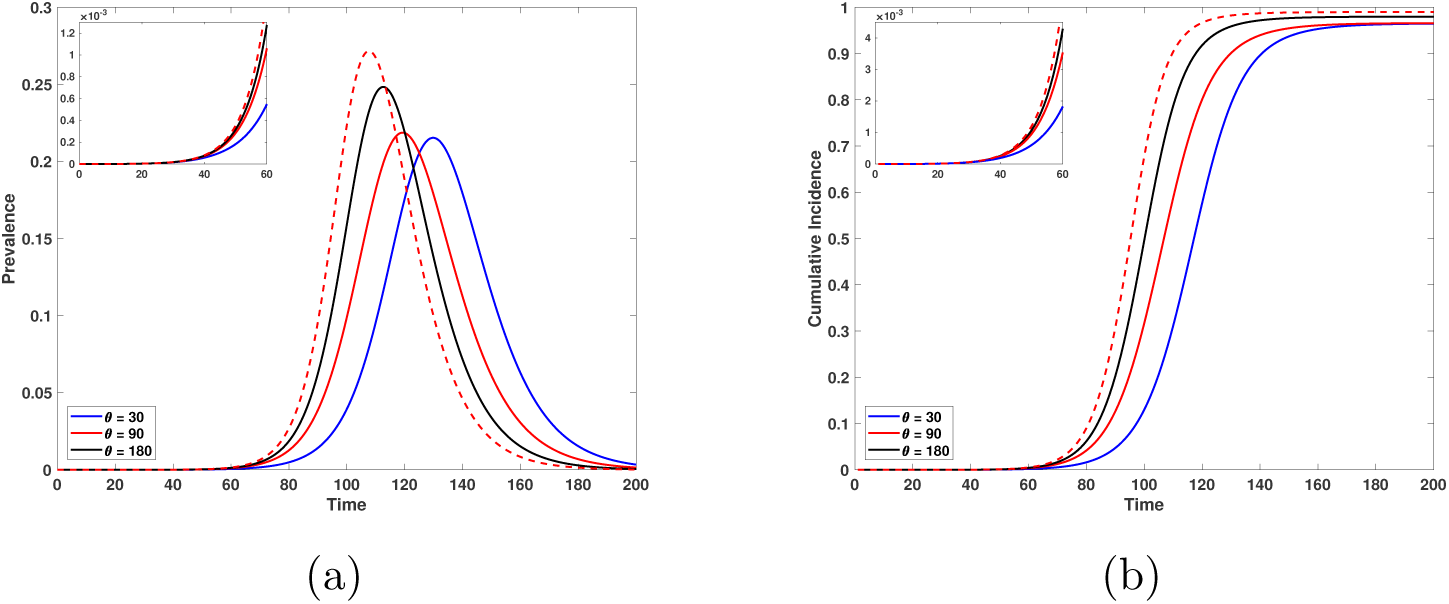
Epidemic curves for a reduction of 25% of the contact rate, the day to start the behavioral change is 15 days after 10 infected are detected and 80% of infectious people are asymptomatic. (*a*) Prevalence. (*b*) Cumulative incidence. Red dashed lines illustrate the epidemic outbreak in the absence of behavioral change. Black, red and blue lines correspond to times obtain the target contact rate reduction of *θ* = 180, 90 and 30 days, respectively. The upper left-hand graph illustrates the initial growth of the epidemic for each case.

**Figure 9:**
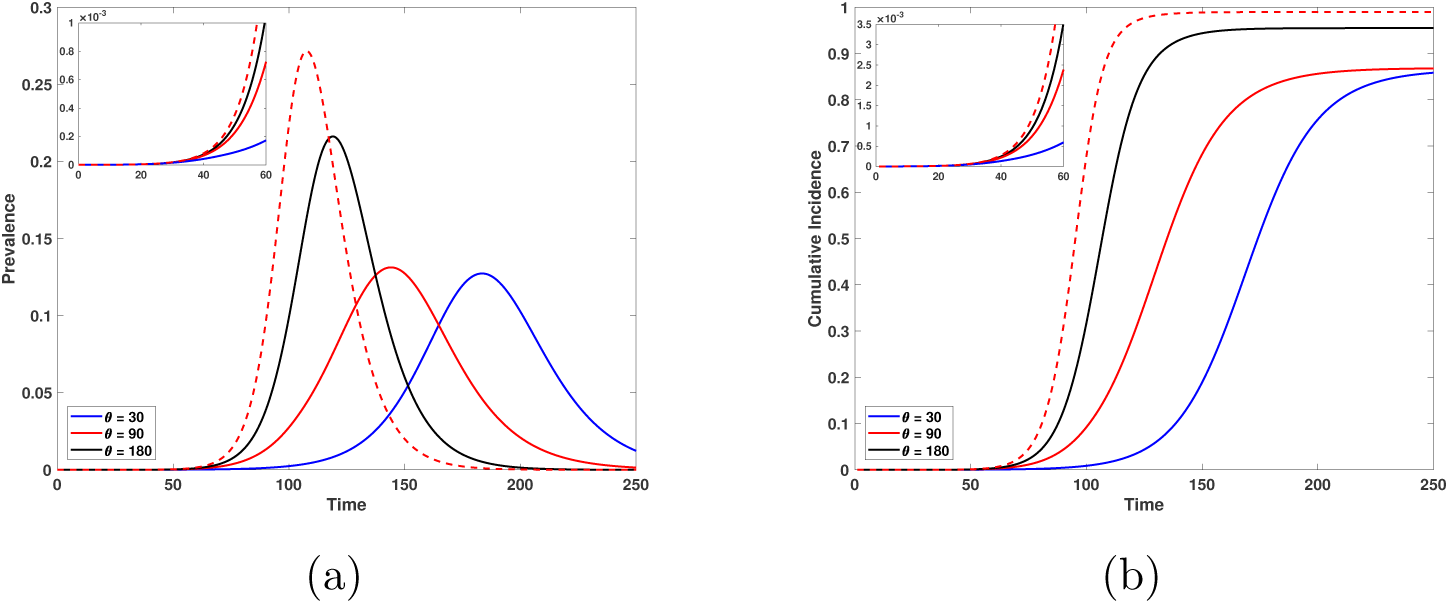
Epidemic curves for a reduction of 50% in the contact rate. The day of start of the behavioral change is 15 days after 10 infected are detected and 80% of infectious individuals are asymptomatic. (*a*) Prevalence. (*b*) cumulative incidence. Red dashed lines depict the behavior of the epidemic outbreak in the absence of behavioral change. Black, red and blue lines correspond to times to obtain the target contact rate reduction of 180, 90 and 30 days, respectively. The upper left-hand graph illustrates the initial growth of the epidemic for each case. Horizontal axis is time in days and the vertical axis is proportions.

Figure 9 shows the behavior of the prevalence (*a*) and cumulative incidence (*b*) when the contact rate is reduced by 50% (*q*_1_ = 0.5). In this situation the relevance of the behavioral change to reduce the incidence is rather clear. Notice that if the time necessary to achieve the contact rate reduction is either *θ* = 30 or *θ* = 90 days (differing for more than two months), the outbreaks are very similar which makes us postulate the existence of a threshold value *θ*^*^ such that the peak incidence is reduced if *θ* < *θ*^*^. Larger learning times do not have an impact on the size of the epidemic.

Figures 10-12 show examples of the behavior of the cumulative incidence (at 180 days after 10 infected are detected which we use as the indicator of initiation of the outbreak in this example) as a function of certain parameter values. Figure 10 shows that if *T*_*θ*_ is closer to the time of detection of the first cases (in this example, when 10 infected people are detected), then the cumulative incidence can be reduced more efficiently and underlines the importance of reducing the effective contact rate quickly. In our example even when the reduction is only of 50%, we see that the final cumulative incidence decreases to low levels. This observation contrasts with what is shown in Figures 8 and 9. We will discuss this in more detail later. On the other hand, Figure 10(b) shows that even when the time it takes to get the contact rate reduction is large (*θ* = 90 days), we can still recover a large reduction in the final cumulative incidence provided we have a quick response to enforce reduced contacts and a large target reduction in the effective contact rate.

**Figure 10:**
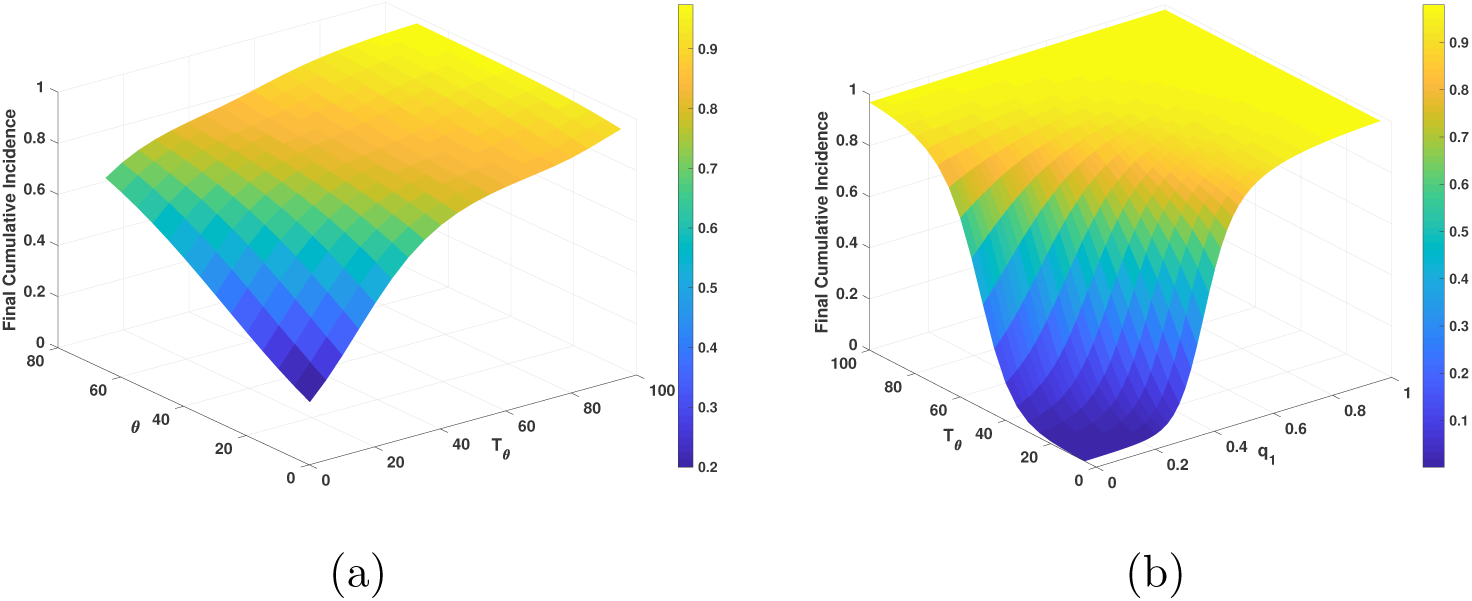
(*a*) Cumulative incidence for varying *θ*, the learning time to achieve the target reduction and *T*_*θ*_, the delay to start behavioral change for a fixed reduction of contact rate of 50%, and (*b*) cumulative incidence for varying *T*_*θ*_, the delay to start behavioral change and *q*_1_ the target proportion of reduction of the contact rate for a fixed time to obtain the target contact rate of 90 days.

Figure 11 shows that when *T*_*θ*_ = 15 days, regardless of the time *θ* it takes to obtain the contact rate reduction, the final cumulative incidence is very low for large reductions in contact rates (*q*_1_ small) or high for low reductions in contact rate (*q*_1_ large).

**Figure 11:**
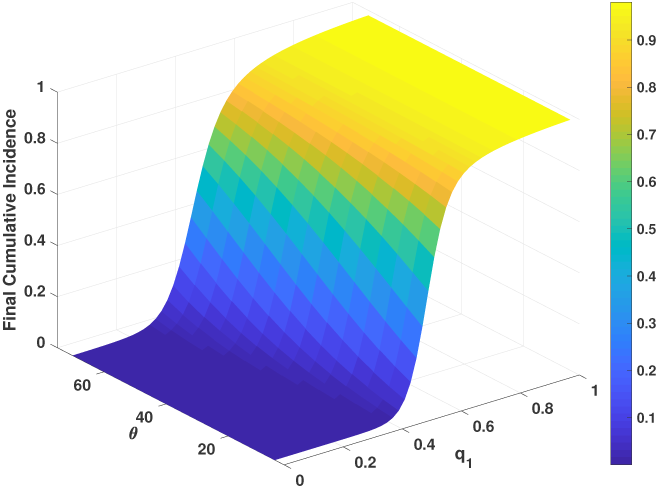
Cumulative incidence when the date to the initial behavioral change is 15 days after 10 cases are detected.

To finish this section, we remark that Figure 10(a) and 12 show the importance of reducing the contact rate for flattening the epidemic curve. On any scenario, a large reduction of the effective contact rate is necessary (Figure 12(a)). Figures 12(b) shows an example where an insufficient reduction of the contact rate (40%), leads to a very weak reduction on the final cumulative incidence.

**Figure 12:**
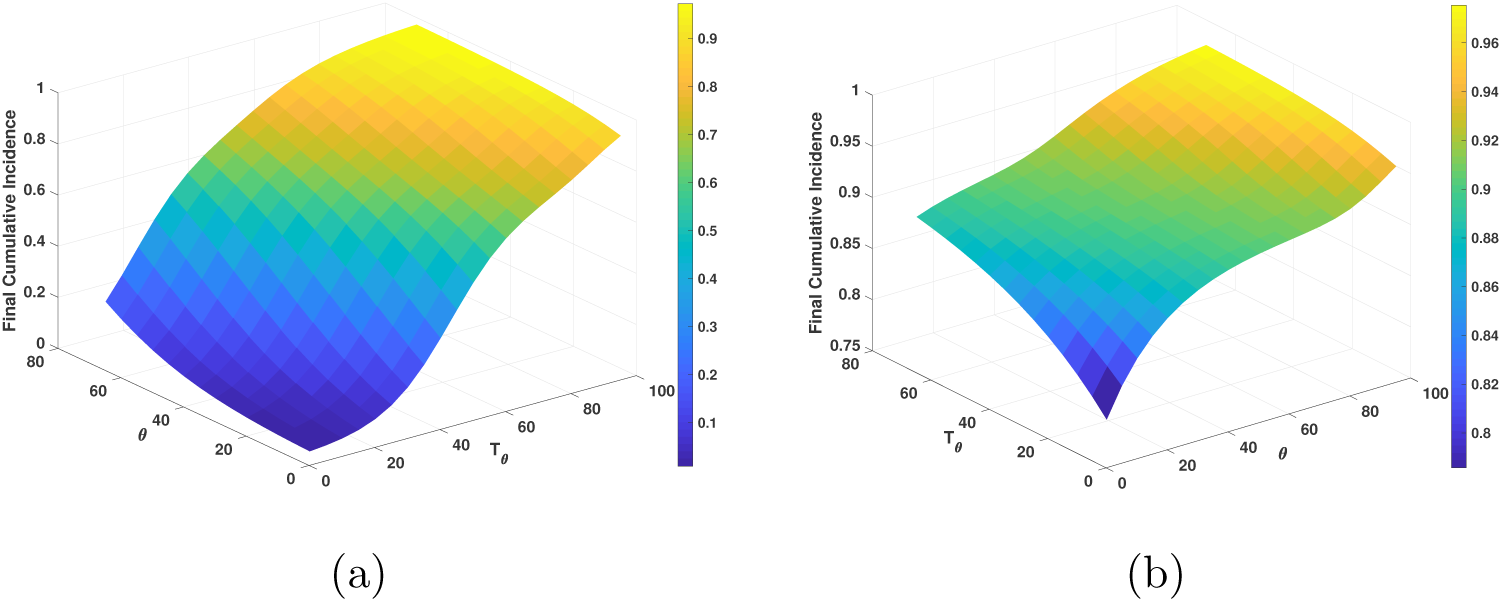
Cumulative incidence when (*a*) the reduction of contact rate is 60%, and (*b*) 40% for varying *θ* and *T*_*θ*_.

## 5 Transmission dynamics under isolation

If the epidemic cannot be contained, social isolation has been implemented in many places of the world, notably in China where, given the extent and strictness of its enforcement, it has apparently been successful. We modify our model to incorporate this containment measure and let *s, e, y*_*a*_, *y*_*s*_ and *r* as usual represent susceptible, exposed, asymptomatic and symptomatic infectious and recovered (immune) individuals in isolation which occurs when a proportion *q* of the general population is put on restricted movement. The population not under isolation (i.e., a fraction 1 − *q* of the total) is denoted by the letters *c*_*s*_, *c*_*e*_, *c*_*y*_*a*, *c*_*y*_*s* and *c*_*r*_ for susceptible, exposed, asymptomatic and symptomatic infectious and recovered (immune) individuals. In this model isolation comprises not only physical isolation, but also the enforcement of sanitary and preventive measures like frequently washing hands, use of face masks, avoiding of public gatherings and so forth.

The actual physical isolation depends on many factors, particularly the length of the incubation period of the virus but many of the preventive measures taken by individuals and groups of individuals can last much longer. The isolation enforcement has the following modeling set up. After the first appearance of the virus a SEIR epidemic starts and develops until time *T*_*θ*_ where quarantine and isolation are enforced and a fraction *q* of individuals in the population goes into isolation. We define *q* to be the efficacy of isolation, e.g., *q* = 0.9 if 90% of the population is isolated.

We then record the number of people in each of the SEIR compartments at *T*_*θ*_, and use these numbers as the initial conditions for the state variables of the same SEIR epidemic but under isolation starting at *T*_*θ*_. The subsystem that escaped or that was not put under isolation we denote by cSEIR.

We also assume that isolation measures are not perfect and individuals escape from isolation regardless of infection status at an exponential average time of 1/*ω* and also that individuals suspend, forget or relax sanitary measures at the rate *ω*. It is also assumed that the isolation time is always larger than the total time of duration of disease comprising the incubation time, 1/*γ*, and the infectiousness time 1/*η*. Therefore 1/*ω* > 1/*γ* + 1/*η*. We further assume that a fraction *ρ* of the exposed individuals develop the asymptomatic disease and that 1 − *ρ* become symptomatic. The contact rates before isolation is enforced are 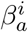 and 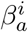 for symptomatic and asymptomatic individuals. Following the ideas set in the previous section we define two time-dependent contact rates for the isolated population that can be consulted in Appendix D.

The natural mortality and recruitment rates respectively are assumed to be negligible during the time of the epidemic. With these definitions the model can is shown in Appendix E. In summary, for *t* < *T*_*θ*_ the infectious disease is the standard SEIR Kermack-MacKendrick model. Once *t* = *T*_*θ*_ is reached, the population is divided into the isolated and non-isolated sections with contact rates (9) and (13) respectively.

### 5.1 Calibration with the Wuhan case

To calibrate our model, in this section we characterize the epidemic in Wuhan, China (epicenter of the first epidemic outbreak) [6]. We introduce an isolation stage and the consequent reduction in the contact rates.

For Wuhan the basic reproduction number has been estimated to be 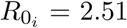 [1, 3, 8, 10] and the parameter baseline

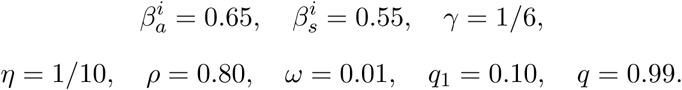

Figure 13 shows the contact rates for the isolated environment. For both rates, the reduction of contact is 90% (*q*_1_ = 0.1), the day of initial isolation is January 26th. We can impose a fast behavioral change because we think that this process lasted a short time (7 days) in Wuhan.

**Figure 13:**
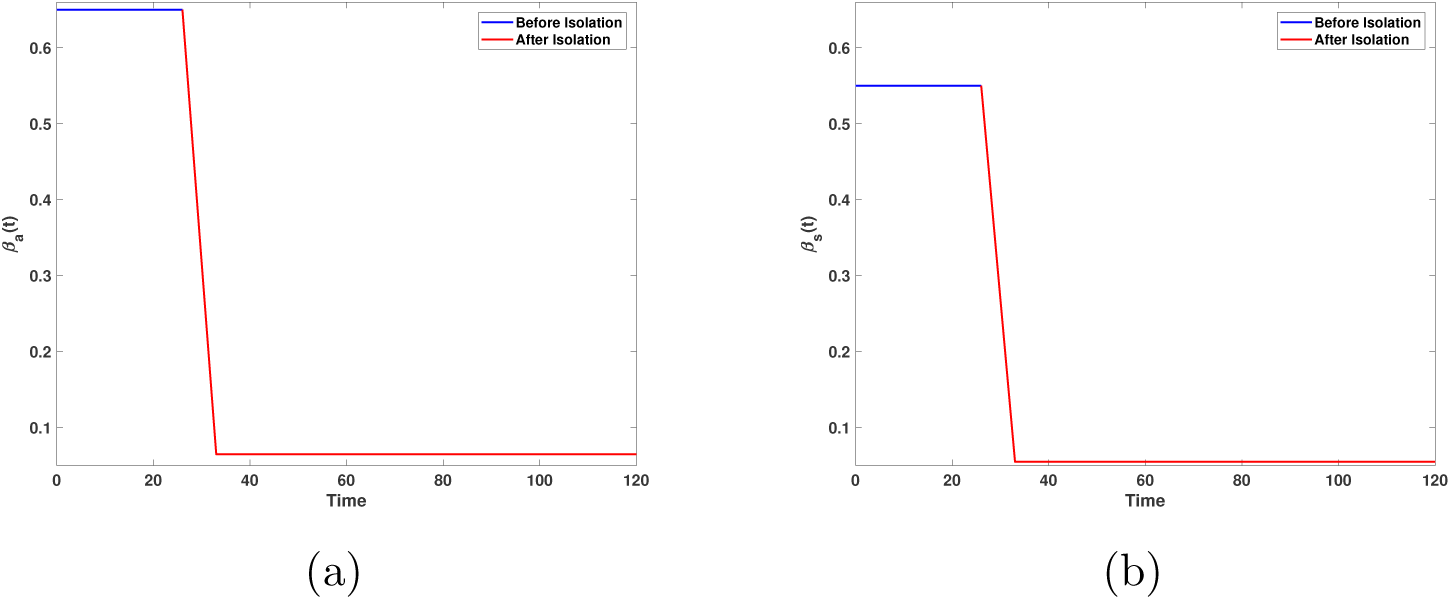
Behavior of time-dependent contact ratesfor a target reduction of contact rate of 90%. The day of initial isolation is January 26th; the time to achieve the target contact rate reduction is 7 days. (*a*) Contact rate for symptomatic and (*b*) symptomatic individuals.

Figure 14 presents the case when the 99% of population is isolated. Observe that the peak incidence is approximately 4 935 individuals and it occurs 2 days after isolation began (January 26th, [20]) Figure 14(b)). Our simulations show that carrying out an isolation process promotes an earlier decrease in incidence. On the other hand, considering a total population equal to 14 million [21] the prevalence at the peak of the epidemic in Wuhan is around 19 000 cases which is a gross underestimate of the true observed value [6]. A better estimate of the at-risk population in Wuhan would, we believe, improve or estimate.

**Figure 14:**
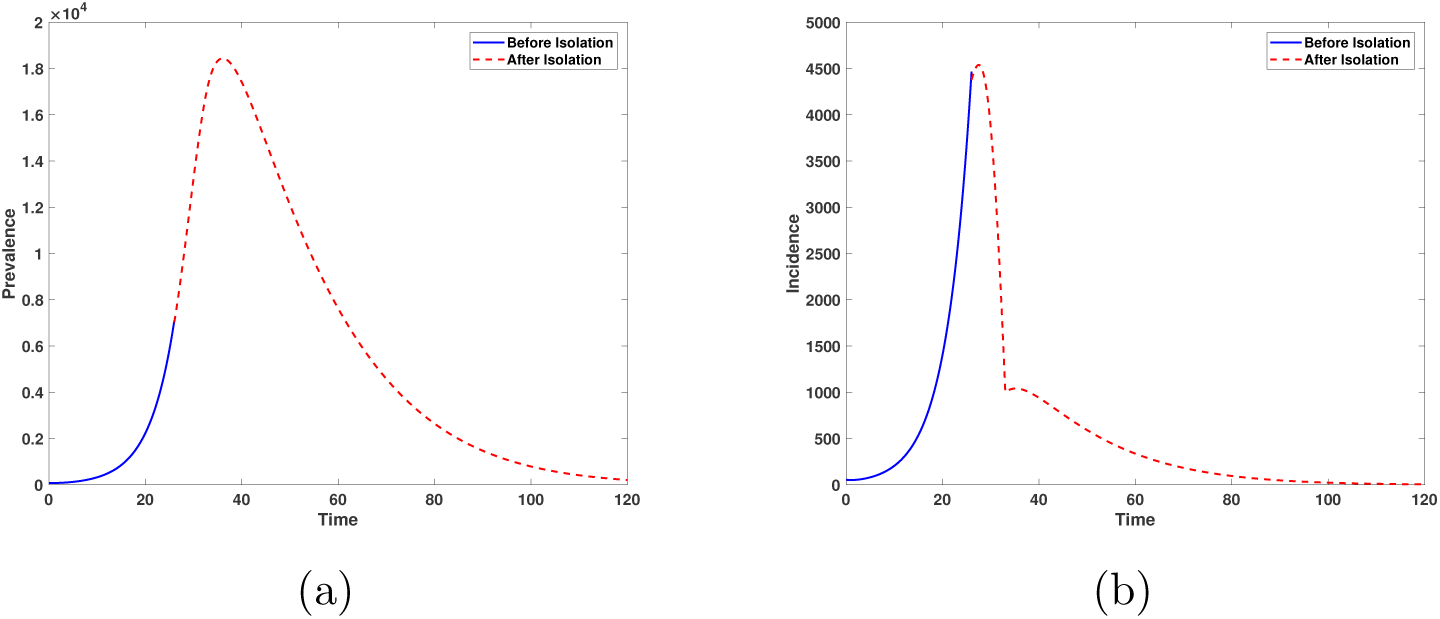
(*a*) Prevalence and (*b*) incidence for the isolated population. The day of initial isolation is January 26th with 99% of the population under isolation; the reduction in contact rate is 90%.

Figure 15 shows a comparison of the estimated cumulative incidence (blue line) versus the data reported by the WHO (red cross) for the Hubei province [20]. Our curves overestimate the data, which may be reasonable due the known under-reporting of cases and to the fact that initially there are fewer cases making it difficult to detect them. Nonetheless we see that when the real epidemic curve begins to stabilize, the difference between both data and model decreases, which can be explained if we accept that, at this stage, there are many more cases and it is easier to identify them. Also under-reporting decreases for the same reason. The black point on the model cumulative incidence (blue curve) indicates the inflection point. The black dots over the red curve mark the range where the inflection point is located in the data.

**Figure 15:**
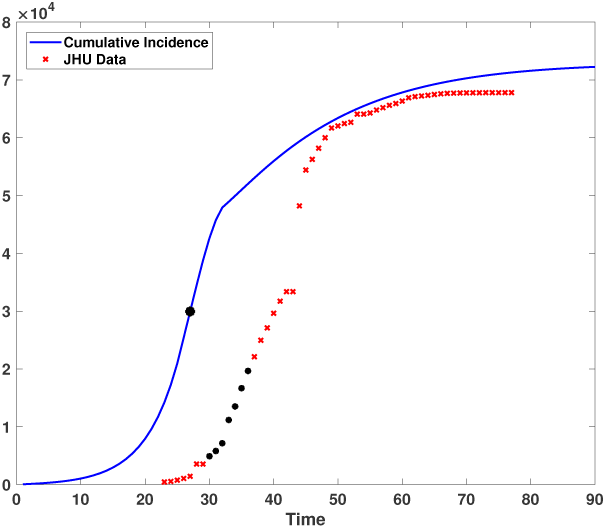
Expected cumulative incidence (blue line) versus Johns Hopkins University data (red cross) for Hubei. Horizontal axis is time in days and the vertical axis is number of individuals.

### 5.2 Understanding the epidemic in Mexico City

The mathematical model in this section is a different variant of the one we applied for Wuhan because Mexico City outbreak started with imported cases. Therefore we retake the model proposed in Section 1 and combine it with the Wuhan model of the previous section to incorporate isolation and behavioral change.

In Mexico air travel has been gradually suspended and at the time of writing (end of March) not all flights have been canceled. This implies that the flow of people from abroad is low but significative and it is arriving into a population that is not isolated but has started measures to reduce the contact rate (social distancing). The model is shown in

Appendix F.

The parameter baseline set is:

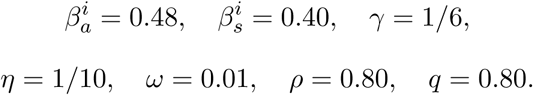

In this section ℛ_0_ = 2.15 consistent with our estimate given in Section 1. Data published by the Mexican Secretary of Health on confirmed cases in the Mexico City and the State of Mexico (that comprise the Greater Metropolitan Area of Mexico City) are 45 (March 20th), 59 (March 21st) and 78 (March 22nd); while our model estimates are equal to 540, 625 and 722, respectively. Our estimates are a reasonable approximation since in many parts of the world the under-reporting generates cases that are approximately a tenth of the true data. Our model simulates infections whereas data shows only confirmed cases.

Below we explore three scenarios that might occur in a large metropolitan that includes variation in the reduction of contact rates, variation in the time it takes to get the contact rate reduction *θ* and different dates for start de isolation *T*_*θ*_. As in Section 1, we consider February 21 as the initial date of infection, Figures 16-18 show the behavior of the outbreak after and before isolation.

**Figure 16:**
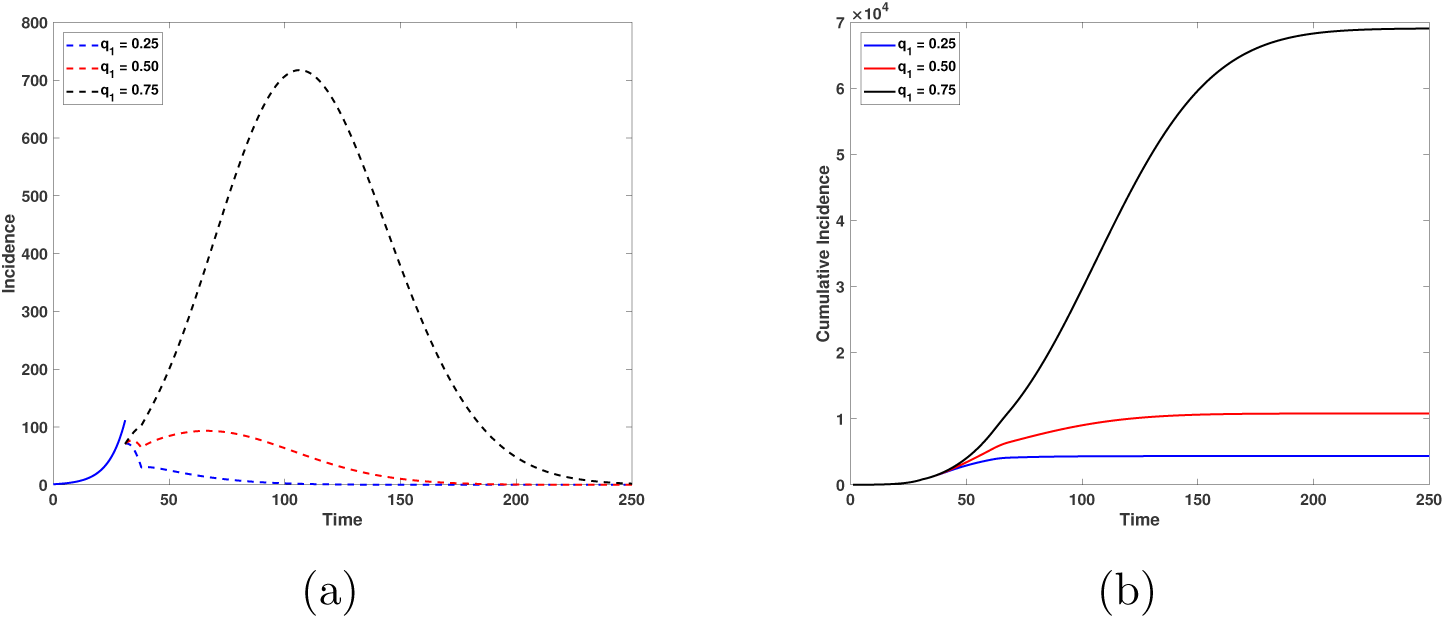
(*a*) Incidence and (*b*) cumulative incidence for the isolated population. The time to obtain the target contact rate reduction is 7 days. Black, red and blue lines correspond to reductions in contact rate of 25%, 50% and 75%, respectively. Horizontal axis is time in days and the vertical axis is number of individuals.

### 5.3 Short time *θ* to reduce the contact rate

On March 23rd authorities of Mexico City ordered the closure of cinemas, theaters, and other public places as a preventive measure. We show in Figure 16 the incidence and cummulative incidence under reductions in contact rates of 25%, 50% and 75%, and for *θ* = 7 days. Our simulations show that for these reductions in contact rates, a reduction of up to 91% in incidence can be obtained. However, if we take the first 35 weeks after the arrival of the first imported case, Figure 16(b) shows that depending on the contact rate reduction, we could reach a similar situation to that of Wuhan with 69 050 total cases, (*q*_1_ = 0.75), or a medium level situation with 10 800 total cases (*q*_1_ = 0.50) or a more manageable situation with 4 351 total cases, (*q*_1_ = 0.25).

### 5.4 Varying *θ*

Figure 17 shows the scenario for different *θ*, the time it takes to get the desired contact rate reduction. In this case isolation also begins on March 23rd and the reduction in contact rates is 60%. We observe that by increasing the value of this parameter, the peak and cumulative incidence will be higher. For example, if the time it takes to reduce the contact rates is *θ* = 7 days, then the cumulative incidence 16 weeks after the arrival of the first imported case will be 6 532, while if the time is *θ* = 15 days, then the incidence rises to 8 909. Finally, if the time it takes to reduce contact rates is 30 days, then the cumulative incidence rises to 14 980. It is important to note that although the value of the cumulative incidence increases, it does not happen with the same intensity as in the case shown in Figure 16.

**Figure 17:**
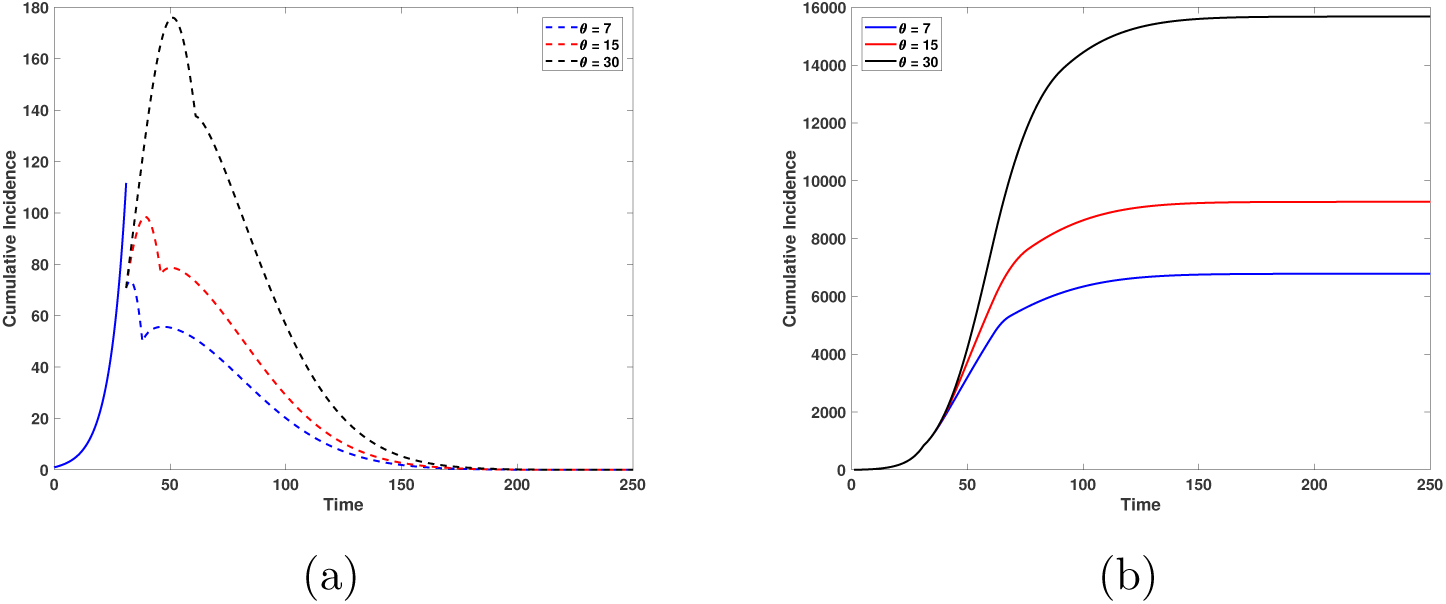
(*a*) Incidence and (*b*) cumulative incidence for the isolated population. The reduction of contact rate is 60%. Blue, red and black lines correspond to times to obtain the target contact rate reduction of 7, 15 and 30 days, respectively. Horizontal axis is time in days and the vertical axis is number of individuals.

### 5.5 Varying the time to start isolation *T*_*θ*_

In our third case we explore variation in the time to start the isolation. Here the time to reduce the contact rates is *θ* = 7 days and the target reduction is 60%. The initial dates of isolation are: March 23rd (blue line), March 30th (red line) and April 6th (black line). The objective of this scenario is to asses what would happen if large cities start isolation and enforce behavioral change several days after this was done in the Mexican capital. Figure 18(a) shows the increase in incidence as the isolation start date increases. Solid lines represent the situation before isolation and the dashed lines represent the situation after the start of isolation. Figure 18(b) shows that if the onset of isolation had occurred one or two weeks after March 23rd, then by week 16, the cumulative incidence would increase by 12 058 and 45 118 new cases, respectively. This scenario underlines the importance of taking contingency measures with due anticipation, otherwise the number of infected people will notoriously increase.

**Figure 18:**
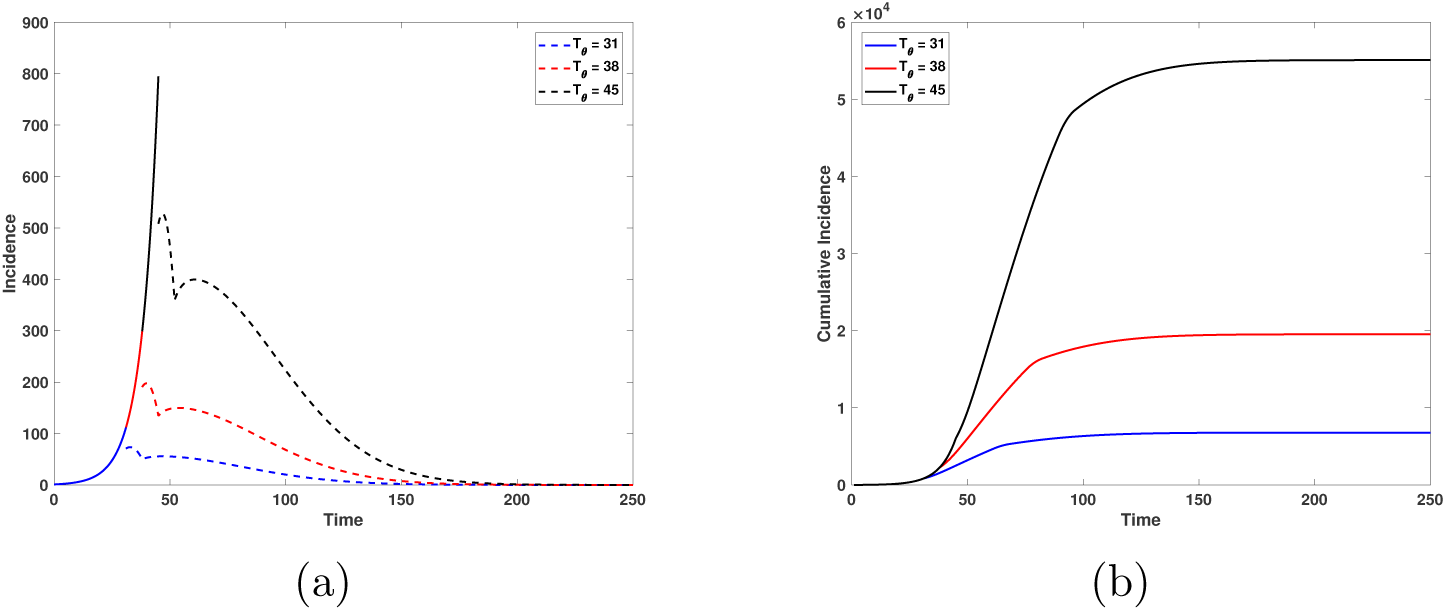
(*a*) Incidence and (*b*) cumulative incidence for the isolated environment. Here we consider that the reduction of contact rate reduction is 60%, and the time it takes to get the contact rate reduction is equal to 7. Blue, red and black lines mean that the dates for start de isolation are 31, 38 and 45 days after the first arrival of an infected individual to Mexico City, respectively. Horizontal axis is time in days and the vertical axis is number of individuals.

## 6 Conclusions

The control, containment, mitigation and final resolution of the coronavirus epidemic requires careful consideration of alternative scenarios as pointed out very clearly in [19]. Models like the ones presented here can help decision-makers to have amore comprehensive understanding of the epidemic and thus a better set of criteria on which to base decisions.

The models presented here are of the same general type that have been published and are being written elsewhere for the coronavirus epidemics. The coronavirus epidemic is an acute respiratory infection highly contagious and with a case fatality rate above of that for influenza. From the modeling point of view the Kermak-McKendrick formalism is the basic and general mathematical tool to understand the epidemic at a population level. The conclusions that we can reach from the study that we have developed in the previous sections are the following:

- Restricting population mobility has been reported as highly effective for the containment of the initial COVID-19 outbreak in China [22,23]. We have attempted to describe the present conditions in Mexico to evaluate the consequences of delaying or advancing the enforcement of isolation and mobility restrictions putting special emphasis on the role of asymptomatic cases since these have been identified as probable important drivers of the disease [13, 24]. At the start of the epidemic in Mexico, Figure 3 shows that the higher the proportion of asymptomatic cases arriving two things happen: the outbreak occurs earlier and also the peak of the outbreak is higher. The role of asymptomatic cases has been under study in the literature particularly as their likely important role in driving the epidemic appears clearer. In our case, their impact is to generate an earlier outbreak and also a higher maximum incidence.
- Figure 4 underlines the relative insensitivity of the peak incidence on the rate of importation of cases provided this rate is relatively low (up to 5 cases per day). Nonetheless, the higher this rate the earlier the outbreak goes into exponential growth (phase 2 according to WHO criteria). We remark that running the simulations that (Figures 3-4) ilustrate for a longer period of time, we can detect the existence of a second epidemic outbreak. With respect to disease spread at a national level in Mexico, the importance of controlling the number of people who arrive infected to the larger airport hubs and bus terminals is of first priority since, not doing it, these can significantly increase the spread of the disease to the other states.
- Behavioral change is one of the main forms to reduce contact rates [19]. However, when dealing with behavioral issues we enter into the psychological realm of individuals and groups of individuals. For our simulations we assume that education or awareness and rational decisions regarding the behavioral change associated with reducing contact rates is the same for asymptomatic and symptomatic individuals although there is some evidence that this might not be the case as one can directly witness. We assume different proportions of these two types of infected individuals. This is an open area of exploration that has not been pursued here. In any case, if asymptomatic individuals or any identifiable portion of the population have more “reckless behavior” on average than other individuals infected or not, then the expected incidence will be higher in any given scenario.
- The policies of containment and mitigation have to be applied quickly, with no hesitation. We observe from our simulations that if the time it takes to obtain the desired contact rate reduction *θ* increases then the reduction rate of both contact rates, for asymptomatic and symptomatic individuals, flattens the curve but more slowly. We also note that in addition to the reduction in cases, as the time to achieve the contact rate reduction increases, the epidemic outbreak starts earlier and is larger. In the examples illustrated above, we see that if the time necessary to achieve the contact rate reduction is either *θ* = 30 or *θ* = 90 days (differing for more than two months), the outbreak sizes are similar which to us indicate the existence of a threshold value *θ*^*^ such that the cumulative incidence is reduced if *θ* < *θ*^*^. In summary large learning times do not have an impact on the size of the epidemic. Figure 10 shows that if *T*_*θ*_ is closer to the time of detection of the first cases (in this example, 10 infected people), then the cumulative incidence can be reduced more efficiently and underlines the importance of reducing the effective contact rate quickly. Figure 10(b) shows that although the time it takes to get the contact rate reduction is high (*θ* = 90 days), we still can obtain a large reduction in the final cumulative incidence provided we have a quick response to enforce reduced contacts and a large target reduction in the effective contact rate. Figure 11 shows that when *T*_*θ*_ = 15 days, regardless of the time *θ* it takes to obtain the contact rate reduction, the final cumulative incidence is very low for large reductions in contact rates (*q*_1_ small) or high for low reductions in contact rate (*q*_1_ large). Figure 10(a) and 12 show the importance of reducing the contact rate for flattening the epidemic curve. On any scenario, a large reduction of the effective contact rate is necessary (Figure 12(a)). Figures 12(b) shows an example where an insufficient reduction of the contact rate (40%), leads to a very weak reduction on the final cumulative incidence.
- Figure 14 shows the case when 99% of population is isolated. We can see that the peak incidence occurs 2 days after isolation began (January 26th) indicating that the isolation process promotes an earlier decrease in incidence. This scenario underlines the importance of taking contingency measures with due anticipation, otherwise the number of cases will notoriously increase.
- Figure 18 highlights the importance of beginning isolation early *T*_*θ*_ = 31 (March 23rd). Our model indicates that delaying action by one or two weeks will let the outbreak grow with a significant increase in cases. Figure 16 shows, however, that isolation is not enough to lower the size of the outbreak but that a significant reduction in transmission rate is also needed. Both control measures have to be applied simultaneously to be efficient in flattening the epidemic curve. We remark that simulations in both, are for a transmission rate reduction achieved in *θ* = 7 days. Figure 17 shows that for longer times, the total number of cases will increase.

All of the latter leaves the message that apart from designing and enforcing control measures, the full participation of the society at large in the fight against this disease is extremely important. It is necessary to reduce the contact rate to a low enough level to be able to contain it, it is necessary to isolate, trace and test individuals. These actions require from everybody to be deeply aware of what should and should not be done to stop the spreading of the disease.

## A Parameter estimation

Let *Y*_*j*_, for *j* = 1, 2, … *n*, be the number of observed cumulative cases at time *t*_*j*_, with *t*_*j*_ given in days. We assume that *Y*_*j*_ follows a Negative Binomial distribution with mean value *C*(*t*_*j*_|*a, r, K*) and dispersion parameter *α*. Here, *C*(*t*_*j*_|*a, r, K*)^1^ is the solution of Richards model presented in (1). Assuming that, given the parameters, the observations *Y*_1_, *Y*_2_, …, *Y*_*n*_ are conditionally independent, then

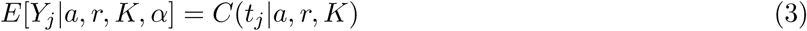

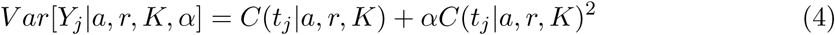

The Negative Binomial distribution allows to control the variability of the data by considering over-dispersion which is common for epidemiological data. If *α* = 0, then we return to the Poisson model which is often used in this context.

Let ***θ*** = (*a*, *r*, *K*, *α*) be the vector of parameters to estimate. The inclusion of the parameter *α*, which is related to the variability of the data, not to the Richards model, is necessary since in practice this variability is unknown. Then, the likelihood function, which represent how likely is to observe the data under the Negative Binomial assumption and Richards model if we knew the parameters, is given by

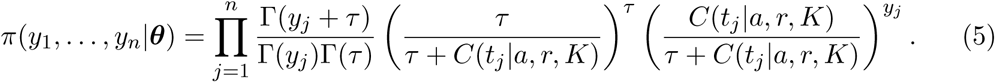

Consider that parameters *a, r, K* and *α* as random variables. Assuming prior independence, the joint prior distribution for vector ***θ*** is

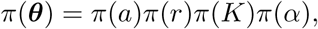

where *π*(*a*) is the probability density function (pdf) of a Uniform(0,1) distribution, *π*(*r*) is the pdf of a Gamma(shape=2, scale=1), *π*(*K*) is the pdf of a Uniform(*K*_min_, *K*_max_), and *π*(*α*) is the pdf of a Gamma(shape=2, scale=0.1). To select the prior for parameter *r*, we consider that previous estimation of *r* are close to 0.3 [15], and a Gamma(2,1) represent a weekly informative prior as it allows for a wide range of values of *r*. Also, there is no available prior information regarding the final size of the outbreak *K*. This is a critical parameter in the model and, in order to avoid bias, we assume a uniform prior over *K*_min_ and *K*_max_. To set these last to values, we consider that the minimum number of confirmed cases is the current number of observed cases *Y* (*t*_*n*_) times nine, i.e. *K*_min_ = *y*_*n*_ ∗ 9. The reasoning behind this number is that it has been suggested that the observed cases are under reported by a factor of nine (REF). To set the upper bound for *K*, we consider a fraction of the total population *K*_max_ = *N* ∗ 0.01, where *N* is the population size of Mexico. This fraction was determined base on the observations of other countries such as Italy where the proportion of infected represents one of the worst case scenarios considering its population size.

Then, the posterior distribution of the parameters of interest is

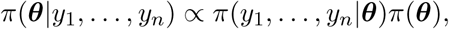

and it does not have an analytical form since the likelihood function depends on the solution of the Richards model, which must be approximated numerically. We analyze the posterior distribution using an MCMC algorithm that does not require tuning called *t-walk* s [25]. This algorithm generates samples from the posterior distribution that can be used to estimate marginal posterior densities, mean, variance, quantiles, etc. We refer the reader to [26] for more details on MCMC methods and to [27] for an introduction to Bayesian inference with differential equations. We run the t-walk for 100,000 iterations, discard the first 25000 and use 1,000 samples to generate estimates of the parameters and short-term predictions for the next 14 days.

## B Case importation model

Equations for the model reviewed in Section 2. See main text for variables and parameter definitions.

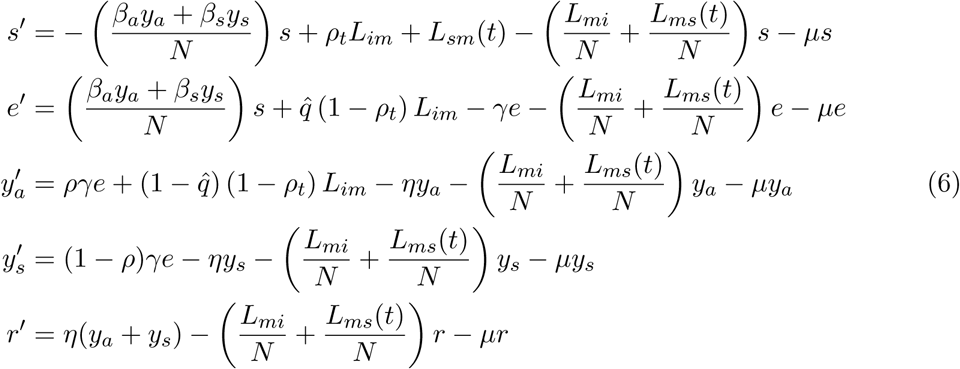

## C National spread model

Equations for the model reviewed in Section 3 that includes transportation. For the analysis we standarize the model variables to work with proportions. We define the variables:

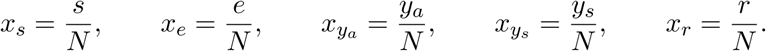

Thus, system (6) becomes

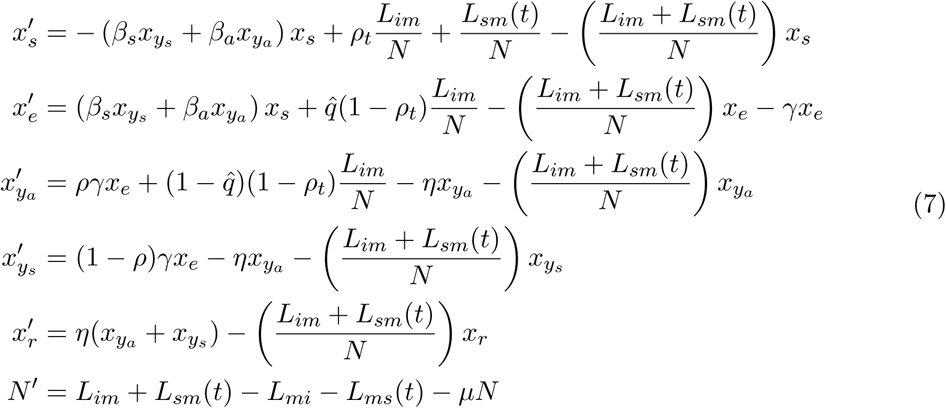

## D Preisolation model

This model sets the migration functios *L*_*im*_ = *Lmi* = *L*_*sm*_ = *L*_*ms*_ = 0 and *µ* = 0. Thus

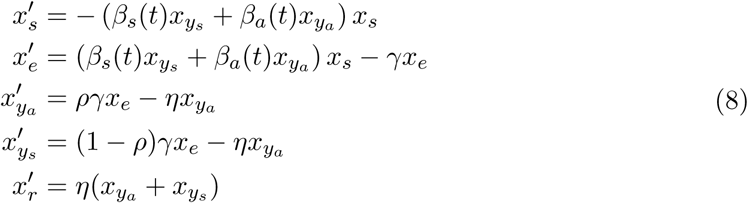

with varying contact rates given as:

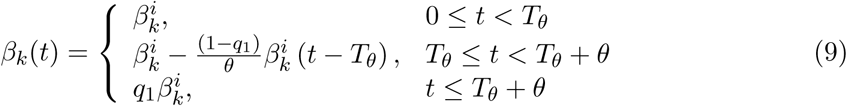

with *k* = *a* or *s* for asymptomatic and symptomatic individuals, respectively.

## E Wuhan model

The model for Wuhan, before containment measures are applied is

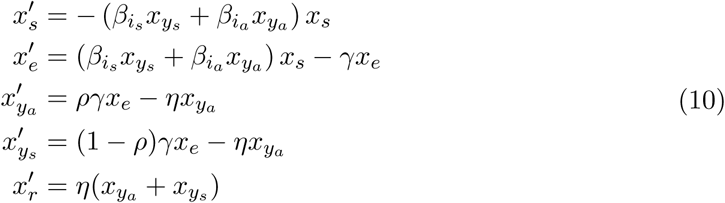

After isolation is enforced, we have a model for the isolated population

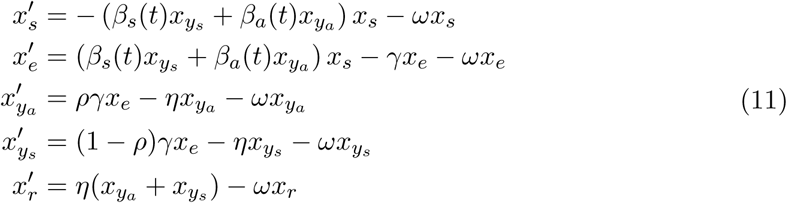

and for the non-isolated population:

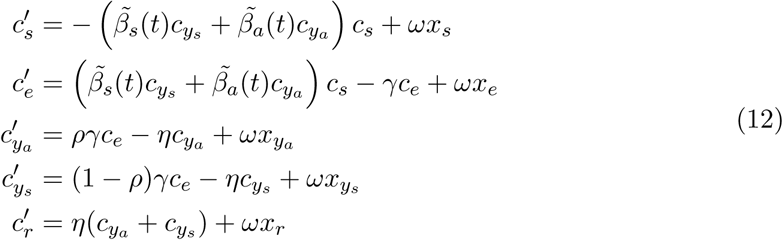

with contact transmission for the non-isolated part of the population:

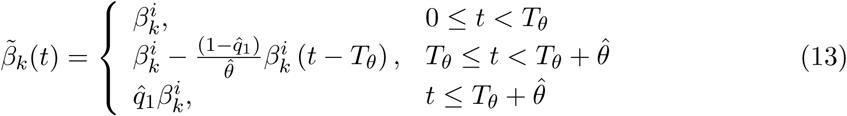

where *k* = *a* or *s* for asymptomatic or symptomatic individuals, respectively. 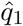 is the desired proportion of reduction of the initial contact rate, e.g., 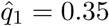 if the contact rate is reduced in 65% at the end of a time interval of length 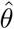.

## F Mexico City model

Dynamics before isolation is given by system 7. After isolation, dynamics for the isolated population is given by

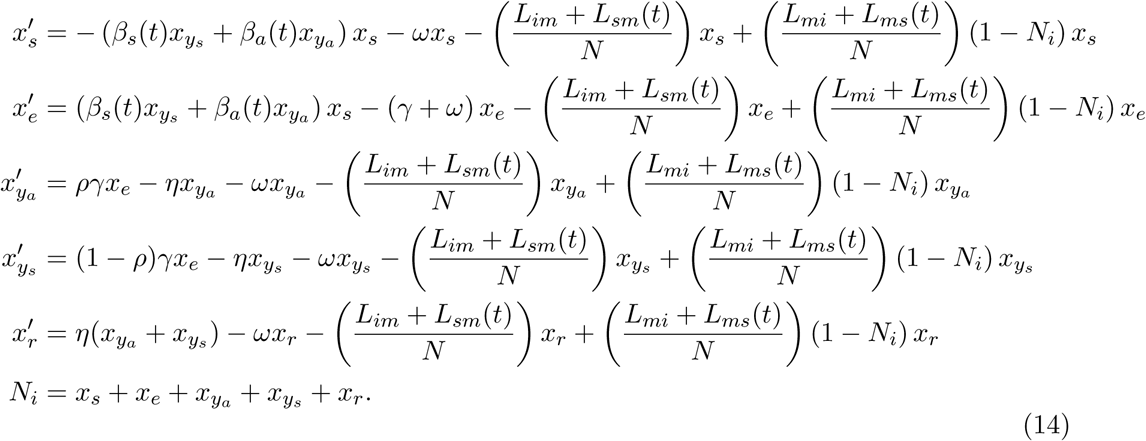

And dynamics for non-isolation is described by:

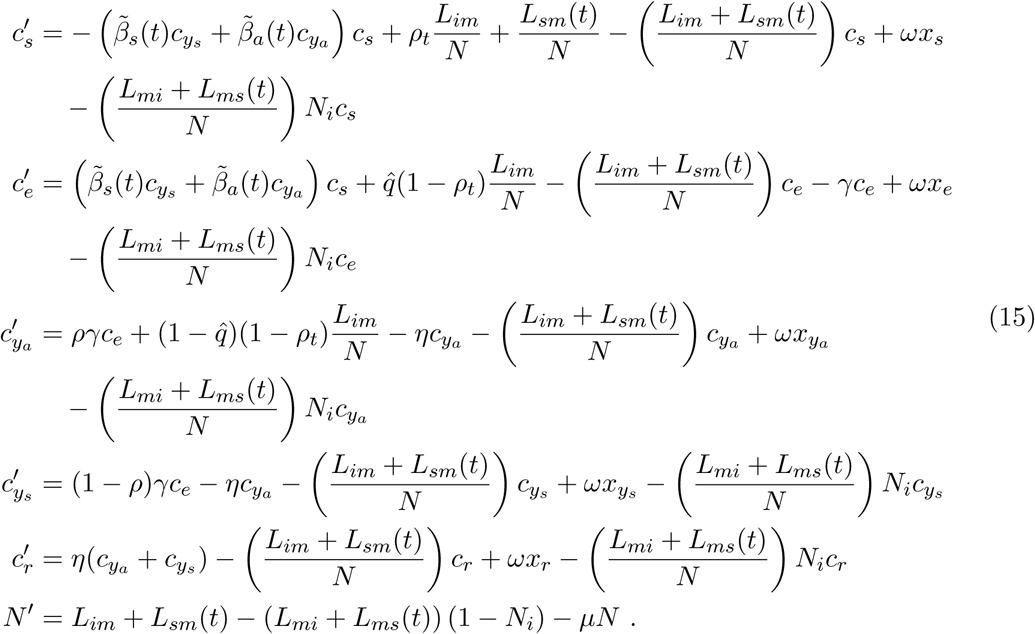

## G Travel data

To estimate the values of *L*_*im*_ and *L*_*mi*_, we calculate the average number of passengers per day arriving/departing from Mexico City from/to abroad taking the total number of passengers for each mean of transportation and divide it by the corresponding number of days. We use the data reported by the Ministry of Communications and Transportation [16].

From Table 4, the values of total air passengers that arrived and departure from Mexico City in February 2020 are 653050 and 633312, respectively. Therefore, *Lim* ≈ 22519 and *Lmi* ≈ 21838.

**Table 4:** Number of passengers arriving/departing from Mexico City (CDMX) from/to abroad in February 2020.

|  | From CDMX | To CDMX |
| --- | --- | --- |
| AMSTERDAM | 12,429 | 13,536 |
| ATLANTA | 17,593 | 18,067 |
| AUSTIN | 1,681 | 1,770 |
| BARCELONA, España | 4,743 | 5,280 |
| BOGOTA | 33,677 | 34,006 |
| BUENOS AIRES | 11,234 | 9,617 |
| CARTAGENA DE INDIAS | 1,844 | 2,104 |
| CHARLOTTE | 3,023 | 3,293 |
| CHICAGO | 20,633 | 20,487 |
| DALLAS-FORT WORTH | 25,420 | 28,138 |
| DENVER | 7,370 | 7,140 |
| DETROIT | 3,370 | 3,233 |
| TOKYO | 8,406 | 9,774 |
| MC ALLEN | 517 | 500 |
| MEDELLIN | 6,299 | 6,298 |
| MIAMI | 30,757 | 32,293 |
| MINNEAPOLIS | 2,940 | 2,702 |
| MONTREAL | 12,262 | 12,797 |
| MUNICH | 3,422 | 4,155 |
| NEWARK | 7,982 | 8,229 |
| NUEVA YORK | 31,012 | 31,675 |
| OAKLAND | 2,593 | 2,494 |
| ORLANDO | 10,781 | 10,927 |
| PANAMA | 19,301 | 20,338 |
| PARIS | 17,895 | 18,885 |
| TORONTO | 18,772 | 18,867 |
| ESTAMBUL (ARNAVUTKOY) | 1,676 | 2,090 |
| FRANKFURT | 8,555 | 9,530 |
| GUATEMALA | 18,343 | 18,684 |
| GUAYAQUIL | 2,876 | 3,547 |
| HOUSTON | 25,892 | 28,416 |
| LA HABANA | 19,320 | 19,773 |
| LAS VEGAS | 14,165 | 14,224 |
| LIMA | 28,768 | 28,046 |
| LONDRES | 8,976 | 9,164 |
| LOS ANGELES | 37,245 | 39,271 |
| MADRID | 28,960 | 29,831 |
| MANAGUA | 1,620 | 1,710 |
| SEOUL | 4,435 | 4,765 |
| PHOENIX | 2,571 | 2,992 |
| QUITO | 6,473 | 7,113 |
| SALT LAKE CITY | 3,939 | 3,906 |
| SAN ANTONIO | 9,426 | 9,925 |
| SAN FRANCISCO | 12,148 | 12,165 |
| SAN JOSE, COSTA RICA | 14,953 | 14,179 |
| SAN PEDRO SULA | 1,654 | 1,882 |
| SAN SALVADOR | 11,471 | 13,149 |
| SANTIAGO DE CHILE | 13,180 | 11,837 |
| SANTO DOMINGO, REP DOM | 2,176 | 2,258 |
| SAO PAULO | 14,805 | 13,796 |
| SEATTLE | 3,454 | 3,656 |
| VANCOUVER | 15,032 | 15,154 |
| WASHINGTON | 2,823 | 2,879 |

The values of *L*_*sm*_ and *L*_*ms*_ outside the Holy Week are estimated similarly to the previous case from data from the Ministry of Communications and Transportation (MCT) and the General Directorate for Tourism of Mexico City [16, 17].

From Table 5, the average number of passengers per day, outside the Holy Week air and terrestrial departing from Mexico City are 31, 173 and 110, 253, respectively. Hence *L*_*ms*_ = 141425. Proceeding in similar manner, the value of *L*_*sm*_ is equal to 141664. Finally, to calculate the *L*_*ms*_ and *L*_*sm*_ values during Holy Week, we increased the above values by 25%. We chose this value because when averaging visits to the Official Sites of the Ministry of Tourism during February and March 2019 [17], and comparing it to the data for the month of Holy Week of the same year, one sees that travel increased by approximately 25% [17]. Therefore, in Holy Week, *L*_*ms*_ ≈ 176781 and *L*_*sm*_ = 177080.

**Table 5:** The Table on the right indicates the number of air passengers arriving/departing from Mexico City (CDMX) from/to some Mexican states in February 2020. The Table on the left shows the number of road passengers in January 2018/2019. ^*^ these values are only rough approximations.

|  | From CDMX | To CDMX |
| --- | --- | --- |
| CANCUN | 180,567 | 186,440 |
| CHIHUAHUA | 29,336 | 28,642 |
| CIUDAD JUAREZ | 24,566 | 23807 |
| GUADALAJARA | 145,432 | 148,335 |
| HERMOSILLO | 31,967 | 30,267 |
| MERIDA | 75,483 | 75,187 |
| MONTERREY | 150,961 | 150,153 |
| OAXACA | 30,978 | 32,398 |
| PUERTO VALLARTA | 42,409 | 44,396 |
| SAN JOS DEL CABO | 36,261 | 35,799 |
| TIJUANA | 87,592 | 81,831 |
| TUXTLA GUTIERREZ | 34,948 | 35,442 |
| VILLAHERMOSA | 33,503 | 34,313 |

|  | From | To* |
| --- | --- | --- |
| North Terminal | 1,062,624 | 1,063,858 |
| East Terminal | 863,604 | 864,621 |
| West Terminal | 1,028,215 | 1,029,324 |
| South Terminal | 463,400 | 464,225 |

## Data Availability

All data is publicly available

## Authors Contributions

Conceptualization: MAAZ, JXVH. Formal Analysis and Investigation: MAAZ, JXVH. Performed statistical analysis: MSC; Wrote and discussed the paper: All authors.

## Acknowledgments

JXVH would like to acknowledge support from grant PAPIIT-UNAM IN115720 and support from the Department of Mathematics of the University of Miami. JXVH also thank Prof. Steve Cantrell and Prof. Chris Costner for their support. We also thank Dr. David Baca Carrasco for his help in the earlier reference search.

## Footnotes

1 We changed the notation with respect to equation 1 to emphasize that the solution of the ODE depends on the parameters of interest.

